# Charting Aging Trajectories of Knee Cartilage Thickness for Early Osteoarthritis Risk Prediction: An MRI Study from the Osteoarthritis Initiative Cohort

**DOI:** 10.1101/2023.09.12.23295398

**Authors:** Tengfei Li, Tianyou Luo, Boqi Chen, Chao Huang, Zhengyang Shen, Zhenlin Xu, Daniel Nissman, Yvonne M. Golightly, Amanda E. Nelson, Marc Niethammer, Hongtu Zhu

**Affiliations:** Department of Radiology, University of North Carolina at Chapel Hill, Chapel Hill, NC, USA; Biomedical Research Imaging Center, School of Medicine, University of North Carolina at Chapel Hill, Chapel Hill, NC, USA; Department of Biostatistics, University of North Carolina at Chapel Hill, Chapel Hill, NC, USA; Department of Computer Science, University of North Carolina at Chapel Hill, Chapel Hill, NC, USA; Department of Statistics, Florida State University, Tallahassee, FL, USA; Department of Epidemiology, University of North Carolina at Chapel Hill, Chapel Hill, NC, USA; Thurston Arthritis Research Center, University of North Carolina at Chapel Hill, Chapel Hill, NC, USA; College of Allied Health Professions, University of Nebraska Medical Center, Omaha, NE, USA

**Author notes:** These authors contributed equally (Tengfei Li, Tianyou Luo).

**Keywords:** Knee osteoarthritis, KLG, knee MRI, local cartilage thickness, aging trajectory, early detection, prognosis

## Abstract

Knee osteoarthritis (OA), a prevalent joint disease in the U.S., poses challenges in terms of predicting of its early progression. Although high-resolution knee magnetic resonance imaging (MRI) facilitates more precise OA diagnosis, the heterogeneous and multifactorial aspects of OA pathology remain significant obstacles for prognosis. MRI-based scoring systems, while standardizing OA assessment, are both time-consuming and labor-intensive. Current AI technologies facilitate knee OA risk scoring and progression prediction, but these often focus on the symptomatic phase of OA, bypassing initial-stage OA prediction. Moreover, their reliance on complex algorithms can hinder clinical interpretation. To this end, we make this effort to construct a computationally efficient, easily-interpretable, and state-of-the-art approach aiding in the radiographic OA (rOA) auto-classification and prediction of the incidence and progression, by contrasting an individual’s cartilage thickness with a similar demographic in the rOA-free cohort. To better visualize, we have developed the toolset for both prediction and local visualization. A movie demonstrating different subtypes of dynamic changes in local centile scores during rOA progression is available at https://tli3.github.io/KneeOA/. Specifically, we constructed age-BMI-dependent reference charts for knee OA cartilage thickness, based on MRI scans from 957 radiographic OA (rOA)-free individuals from the Osteoarthritis Initiative cohort. Then we extracted local and global centiles by contrasting an individual’s cartilage thickness to the rOA-free cohort with a similar age and BMI. Using traditional boosting approaches with our centile-based features, we obtain rOA classification of KLG ≤ 1 versus KLG = 2 (AUC = *0.95*, F1 = *0.89*), KLG ≤ 1 versus KLG ≥ 2 (AUC = *0.90*, F1 = *0.82*) and prediction of KLG2 progression (AUC = *0.98*, F1 = *0.94*), rOA incidence (KLG increasing from < *2* to ≥ *2*; AUC = *0.81*, F1 = *0.69*) and rOA initial transition (KLG from 0 to 1; AUC = *0.64*, F1 = *0.65*) within a future 48-month period. Such performance in classifying KLG ≥ 2 matches that of deep learning methods in recent literature. Furthermore, its clinical interpretation suggests that cartilage changes, such as thickening in lateral femoral and anterior femoral regions and thinning in lateral tibial regions, may serve as indicators for prediction of rOA incidence and early progression. Meanwhile, cartilage thickening in the posterior medial and posterior lateral femoral regions, coupled with a reduction in the central medial femoral region, may signify initial phases of rOA transition.

## Introduction

Knee osteoarthritis (OA), a common chronic disease characterized in part by cartilage degeneration, inflammation, and pain (1, 2), presents a substantial global health and economic burden. Its global prevalence reached 364.6 million in 2019 (3), with 86.7 million new adult cases reported in 2020 (4). Nearly 30% of individuals above 45 demonstrate signs of knee radiographic OA (rOA), with around half reporting symptoms, and significantly more severe among women (5-7). Other health implications of knee OA include frequent comorbidities (8). Economically, OA and related disorders entail substantial total costs of approximately $486.4 billion, with $136.8 billion directly attributable to OA (9), and a lifetime cost of $15,000 per patient (6, 10). Given the aging population and rising obesity rates, these statistics are likely to be further amplified in the future, underscoring the importance of knee OA management.

While knee OA is prevalent, its complex etiology involving various joint structures like cartilage, bone, synovium, ligaments, periarticular fat, meniscus, muscle, biomechanical influences, immune response variations (6, 11, 12), and their intricate interplay (13), complicates the prognosis and treatment of knee OA. Existing management strategies range from patient education (14), exercise (15), and weight management (16), to pharmacological interventions using nonsteroidal anti-inflammatories (17) and pain alleviation drugs (18). However, in advanced cases, total knee replacement (TKR) surgeries, although invasive and costly, become necessary. Timely diagnosis can decelerate OA progression, mitigate the need for TKR and curtail long-term medication use and associated adverse events, underscoring the need for effective early interventions for knee OA.

Diagnosing early OA, when typical signs and symptoms of advanced OA are not apparent, is challenging. Traditional radiographs primarily detect changes in bone structure, such as joint space narrowing or osteophyte formation, common in later stages (19). In early OA, tissue-related changes such as cartilage or synovial damage, which may not be visible on radiographs, can occur (20, 21). On the other hand, magnetic resonance imaging (MRI), which visualizes all joint tissues, can detect a diverse array of pathological changes, such as early cartilage damage, bone marrow lesions, synovitis, or changes in the menisci, ligaments, or tendons, that cannot be captured by radiographs (22). Through various modalities, OA can be assessed and classified by radiographic OA (rOA), MRI-identified OA (MOA) and histopathological OA (hOA) (20). The widely accepted Kellgren-Lawrence Grading (KLG) (23) system has long served as the rOA standard across clinical practice and research. This system classifies OA severity on a scale from 0 (rOA-free) to 4 (severe rOA) using radiographs. Despite its efficacy in detecting moderate (grade 3) to severe rOA, the KLG system’s sensitivity is limited in accurately identifying early rOA stages, such as the grade 2 (24), and grade 1 (25). Enhancements in early OA detection have been driven by the development of MRI scoring systems such as the Magnetic Resonance Imaging Osteoarthritis Knee Score (MOAKS) (19), the Knee Osteoarthritis Scoring System (KOSS) (26), the Boston Leeds Osteoarthritis Knee Score (BLOKS) (27), and the Whole-Organ Magnetic Resonance Imaging Score (WORMS) (28). These systems capture early tissue changes more sensitively and are crucial for defining MOA. For instance, the European Society of Sports Traumatology, Knee Surgery and Arthroscopy (ESSKA) describes (29) early MOA using WORMS grades 3–6 for cartilage morphology Scores, 2-3 for bone marrow lesions, ≥ 2 for cartilage regional loss, and ≥ 3 for meniscal tears. From a histopathological perspective, early OA is determined (20) when ≤ 50% of the cartilage thickness is involved, as per the macroscopic International Cartilage Repair Society (ICRS) classification. Considering these standards, early knee OA is classified (29) based on, a KLG of 0, 1, or 2 in conjunction with knee pain, with additional evidence such as arthroscopic findings of cartilage lesions, MRI evidence of articular cartilage or meniscal degeneration, or subchondral bone marrow lesions.

Over recent decades, a plethora of literature has been dedicated to extracting MRI-based scores or features to predict the future incidence of rOA. This includes the use of MOAKS (30, 31) and WORMS (32), and MRI bone shape features (33), among others, to predict incident rOA 12 months to 48 months prior. These studies propose that MRI changes in bone can presage the early development of rOA and knee pain. However, these methods have notable limitations. Foremost, they are both labor-intensive and time-consuming, necessitating a considerable investment of time and requiring significant clinical expertise for manual scoring or landmark labeling. Moreover, existing MRI scoring systems primarily focus on cartilage loss and lesions, while potentially overlooking the early stage swelling or thickening of the cartilage. It has been suggested (34) that the pathogenesis of OA might not be unidirectional, involving solely cartilage loss. On the contrary, some studies have reported that cartilage can also thicken in the early stages of OA, a phenomenon observed in both animal studies (35-39) and humans (34, 40-44) with early OA. Cartilage thickening in early OA stages, possibly resulting from water diffusion, inflammation, or an early reparative response (34), is not yet fully understood. These findings highlight the need for refinement and improvement of these scoring systems for better early detection of rOA.

On the other hand, despite the advancements in machine learning (ML) and deep learning (DL) for automating OA classification and prediction, significant challenges in this area remain. These technologies have been deployed for a broad range of applications (45, 46), including OA classification (47-57) and prediction (58-65), joint space narrowing (JSN) prediction (66-68), pain symptom classification (69-71) and prediction (72), cartilage lesion detection (73) and TKR incidence prediction (74-79) using MRI or radiograph data. However, the complex inner workings of these artificial intelligence (AI) algorithms often remain opaque, leading to limited interpretability (45). Efforts towards improving the transparency of DL-based models persist (80, 81), yet it remains a major challenge. Additionally, the reliance on the KLG system to classify the severity of rOA presents another limitation. The KLG system, while widely accepted, may not be well suited to early-stage disease detection. These models often focus on detecting and predicting OA progression in its later, symptomatic phases, ignoring OA at the early stages. Models capable of detecting and predicting the earliest stages of OA are needed.

In this study, we aim to predict the early rOA progression, including the potential transition from normal to rOA initial onset and from the initial stage to mild stage. To accomplish this, we first utilized MRI data to establish reference charts for rOA-free knee thickness. Our training data for these reference charts comprised 5,045 scans from 957 participants without rOA (KLG = 0), aged between 45 and 79, who were enrolled in the publicly available Osteoarthritis Initiative (OAI) dataset between 2004 and 2006 (82) (https://nda.nih.gov/oai/). We extracted local cartilage thickness (LCT) maps from knee MRIs using automatic segmentation and registration techniques (83, 84). Next, we constructed the aging charts for LCT among the rOA-free participants, using a nonlinear generalized additive modeling framework with mixed effects (GAMM), accounting for confounding factors such as age, sex, race, baseline height, and body mass index (BMI). We then evaluated centile scores for each individual based on their ranking among peers with similar demographics who were free from rOA. Using these centile score features, we conducted three predictive analyses. (i) Generated a MRI-based rOA auto-classification system with high accuracy, in classifying the early rOA phases into KLG0-1, and KLG2 with and without progression in 48 months. (ii) Predicted the early rOA transition (from KLG = 1 to 2). (iii) Predicted the rOA initial transition (from KLG = 0 to 1). In particular, we may have the following contributions to the field.

1. We developed comprehensive knee cartilage thickness growth reference charts, considering complex nonlinear relationships among age, BMI, and their interactions. Although the effects of sex, age, and BMI have been widely studied in previous literature (6), their complex nonlinear relationships have rarely been investigated. By hypothesizing that distinct cartilage thickness growth curves exist across different knee-OA groups, this approach could distinguish rOA phases effectively.
2. We introduced an automated, easily interpretable and local MRI-based scoring system, based on rankings of cartilage thickness growth trajectories among rOA-free participants. This system could facilitate KLG ≤ 1 versus KLG = 2 classification (AUC *= 0.95*, F1 *= 0.89*), KLG ≤ 1 versus KLG ≥ 2 classification (AUC *= 0.90*, F1 *= 0.82*) and prediction of KLG2 progression (AUC *= 0.98*, F1 *= 0.94*), incidence rOA transition (AUC = 0.81, F1 = 0.69) and rOA initial transition (AUC = 0.64, F1 = 0.65).
3. Our findings supported the notion that cartilage thickening is an important biomarker in early rOA transition (34). We reported key biomarkers for the rOA initial transition, incidence-rOA transition and rOA early progression, such as anterior femoral cartilage thickening and lateral tibial cartilage thinning during the progression from KLG2 to KLG3.

We outlined our workflow and analysis process in **Figure 1**. Our work may offer valuable insights into OA by establishing a standard reference for cartilage aging comparison, and the early detection of OA transition.

**Fig. 1.**
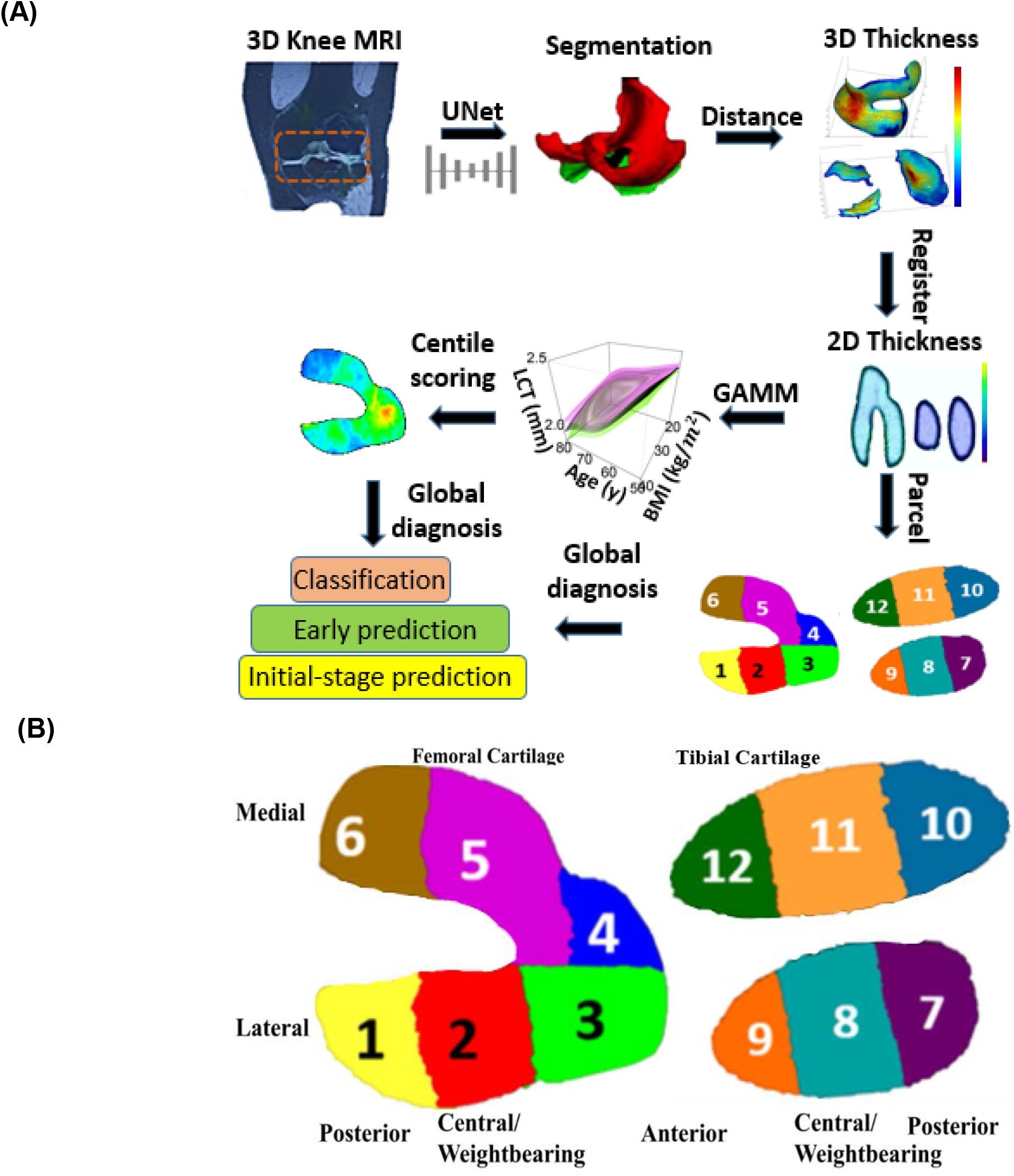
Overview of the study design. **(A)** Overall workflow of the study. We took the 3D MRI from the OAI dataset as an input and generated the 2D (femoral and tibial) cartilage thickness map on the standard space. We fit aging trajectories at each spatial location and build the MRI centile scoring system. The centile scores were used to predict scans scans with future rOA progression. **(B)** Parcellations of the knee cartilage. Regions of interest (ROIs) 1-6 correspond to femoral cartilage; ROIs 7-12 correspond to tibial cartilage. 1: Lateral posterior femur (LPF); 2: Lateral central femur (LCF); 3: Lateral anterior femur (LAF); 4: Medial anterior femur (MAF); 5: Medial central femur (MCF); 6: Medial posterior femur (MPF); 7: Lateral posterior tibia (LPT); 8: Lateral central tibia (LCeT); 9: Lateral anterior tibia (LAT); 10: Medial posterior tibia (MPT); 11: Medial central tibia (MCT); 12: Medial anterior tibia (MAT).

## Results

Following prior literature (85-87), we extended specific terminology based on the KL grades.

1. Knees. Knees were classified into KLG0, KLG1, KLG2, KLG3 and KLG4 according to KLG = 0, 1, 2, 3, 4, respectively. Scans with KLG ≥ 2 were designated as definite-rOA knees.
2. Participants. We separated participants into two groups, the **KLG0-1-stable** (KLG ≤ 1 across all visits within 96 months for both knees) and **rOA-prevalence** cases (KLG attains 2 within 96 months).

The KLG0-1-stable group included three subgroups,

i. the **rOA-free** (KLG = 0 across all visits for both knees),
ii. the **KLG1-stable** (KLG = 1 across all visits)
iii. the **KLG0-1-transition** (baseline KLG = 0 which transits to KLG = 1).

The rOA-prevalence group included two subgroups,

i. the **rOA-incidence** (baseline KLG ≤ 1 which transits into KLG ≥ 2), and
ii. the **KLG2-4-progressive** (KLG ≥ 2 across all visits),

where the rOA-incidence subgroup includes the following subsets,

iii) the **KLG1-progressive** (baseline KLG = 1 and then to KLG ≥ 2),
iv) the **KLG0-progressive** (baseline KLG = 0 and then to KLG ≥ 2),

and both the rOA-incidence and KLG2-4-progressive subgroups included,

v) the **KLG2-3-transition** (KLG = 2 at one visit which transit into KLG = 3 in later phases),
vi) the **KLG2-stable** (KLG = 2 at one visit and ≤ 2 until the last visit).

Classifications for other subpopulations were applied to left and right knees, separately. Furthermore, we referred to the rOA initial transition as the KLG transition from 0 to 1, and termed the incidence rOA transition as the KLG shift from less than 2 to 2. In **Table 1**, we offered descriptive statistics for 9 primary subpopulations, which included sample size, age, sex, BMI, race, and height distributions from the participants’ initial visits.

**Table 1.** Descriptive statistics for 9 subpopulations in OAI. Those includes sample size of participants, number of visits, sample size for females and whites, and BMI, height and age at the baseline visits, of the KLG0-1-stable (KLG ≤ 1 across all visits), rOA-prevalence (all KLG >1 cases), rOA-free (KLG = 0 across all visits), rOA-incidence, KLG0-1-transition, KLG1-progressive, KLG2-3 transition and KLG2-stable participants. Statistics are based on the right knees.

|  | KLG0-1-stable |  | rOA-prevalence |  |
| --- | --- | --- | --- | --- |
|  | Female | Male | Female | Male |
| **Number of Participants** (visits) | 769 (4131) | 593 (3339) | 1252 (5602) | 792 (3716) |
| **Sex %** | 56.5% | 43.5% | 61.3% | 38.7% |
| **Race for white (%)** | 665 (86.5%) | 525 (88.5%) | 917 (73.2%) | 670 (84.6%) |
| **Baseline BMI ± sd** (kg/m<sup>2</sup>) | 26.2±4.6 | 27.9±4.1 | 29.9±5.3 | 29.7±4.1 |
| **Baseline height ± sd** (cm) | 162.2±5.9 | 176.2±6.4 | 162.8±6.2 | 176.3±6.5 |
| **Baseline age ± sd** (year old) | 59.8±9.1 | 58.5±9.2 | 62.5±8.8 | 62.4±9.2 |
|  | rOA-free |  | rOA-incidence |  |
|  | Female | Male | Female | Male |
| **Number of Participants** (visits) | 542 (2799) | 415 (2246) | 134 (763) | 79 (451) |
| **Sex %** | 56.6% | 43.4% | 62.9% | 37.1% |
| **Race for white (%)** | 467 (86.2%) | 361 (87.0%) | 124 (80.5%) | 82 (92.1%) |
| **Baseline BMI ± sd** (kg/m<sup>2</sup>) | 25.8±4.5 | 27.7±4.1 | 29.6±5.2 | 29.4±3.9 |
| **Baseline height ± sd** (cm) | 162.3±5.9 | 176.3±6.5 | 163.1±6.6 | 176.5±7.1 |
| **Baseline age ± sd** (year old) | 59.3±9.2 | 57.4±8.9 | 60.3±8.8 | 61.4±8.7 |
|  | KLG0-1-transition |  | KLG1-progressive |  |
|  | Female | Male | Female | Male |
| **Number of Participants** (visits) | 27 (155) | 19 (99) | 94 (537) | 49 (271) |
| **Sex %** | 58.7% | 41.3% | 65.7% | 34.3% |
| **Race for white (%)** | 25 (92.6%) | 17 (89.5%) | 77 (81.9%) | 46 (93.9%) |
| **Baseline BMI ± sd** (kg/m<sup>2</sup>) | 24.7±3.7 | 28.5±5.8 | 29.9±4.5 | 29.7±4.2 |
| **Baseline height ± sd** (cm) | 163.6±6.9 | 176.1±5.6 | 163.2±6.8 | 177.5±7.5 |
| **Baseline age ± sd** (year old) | 58.2±7.7 | 53.7±7.7 | 60.6±8.9 | 60.7±8.6 |
|  | KLG2-4-progressive |  | KLG2-3-transition |  |
|  | Female | Male | Female | Male |
| **Number of Participants** (visits) | 1118 (5313) | 713 (3546) | 126 (571) | 53 (251) |
| **Sex %** | 61.1% | 39.9% | 70.4% | 29.6% |
| **Race for white (%)** | 807 (72.2%) | 597 (83.7%) | 92 (73.0%) | 44 (83.0%) |
| **Baseline BMI ± sd** (kg/m<sup>2</sup>) | 30.0±5.4 | 29.7±4.1 | 30.7±5.1 | 29.7±4.3 |
| **Baseline height ± sd** (cm) | 162.8±6.1 | 176.3±6.5 | 163.1±7.0 | 176.5±7.0 |
| **Baseline age ± sd** (year old) | 62.9±8.8 | 62.4±9.3 | 61.8±7.9 | 60.9±9.3 |
|  | KLG2-stable |  |  |  |
|  | Female | Male |  |  |
| **Number of Participants** (visits) | 495 (2670) | 315 (1719) |  |  |
| **Sex %** | 61.1% | 38.9% |  |  |
| **Race for white (%)** | 368 (74.3%) | 270 (85.7%) |  |  |
| **Baseline BMI ± sd** (kg/m<sup>2</sup>) | 29.3±5.2 | 29.4±3.9 | | |
| **Baseline height ± sd** (cm) | 163.1±5.8 | 176.0±6.4 | | |
| **Baseline age ± sd** (year old) | 61.2±8.7 | 61.0±9.4 | | |

### Reference cartilage thickness charts highlight thickness growth patterns

We modeled the sex-specific trajectories of cartilage thickness with respect to age and BMI, adjusted for race and height (detailed methods are available in the *Methods* Section). **Figure 2A** provides reference charts depicting the non-linear interplay among the BMI, age, and average cartilage thickness in the tibial and femoral regions of the right knee, stratified by sex. These charts elucidate complex nonlinear associations through 3D perspective and 2D contour plots, with the corresponding confidence interval surfaces. Detailed reference charts for 12 regions of interest (ROIs) in both left and right knees are presented in the **Supplementary Figures S1** and **S2. Figure 2B** presents the effect size and statistical significance of sex, race, height, BMI, age-sex interaction, and sex-BMI interaction across all the regions.

**Fig. 2.**
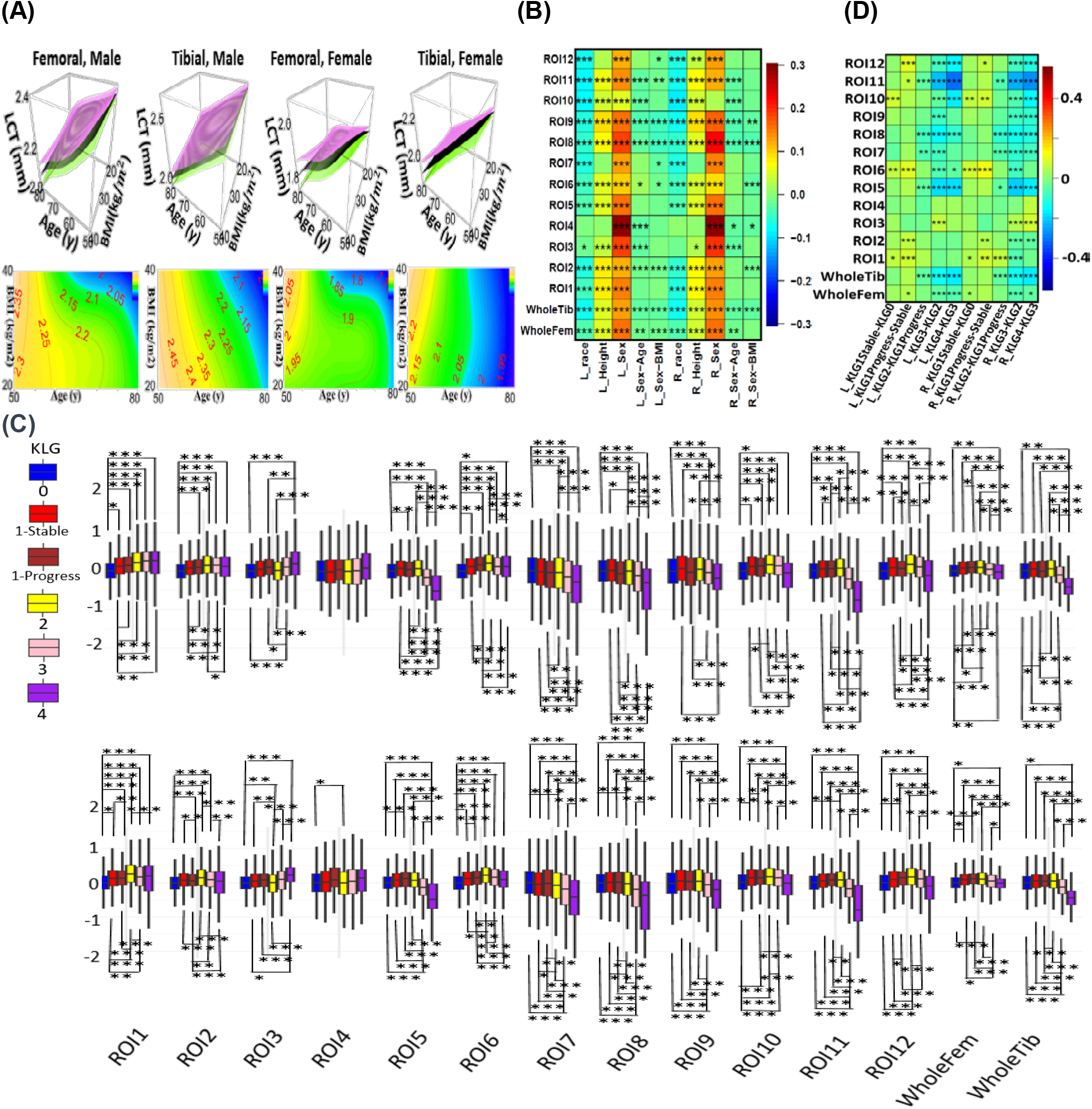
Results of association analyses. **(A)** Overview of the trajectories of cartilage thicknesses in terms of BMI and age on the right knee, with both the 3D perspective view (row 1) and 2D contour plots (row 2) for the overall tibial and femoral regions among males and females, respectively. *95%* confidence surfaces are shown in purple (upper) and green (lower) colors. **(B)** Heatmap to show the effect sizes (standardized coefficients) of race, height, sex, sex*age, sex*BMI on cartilage thicknesses in tibial, femoral and *12* regions of interest (ROIs) for left (column 1-5) and right (column 6-10) knees. **(C)** Boxplots depict cartilage thickness (adjusted for confounders) for left (upper) and right (lower) knees. Different colors represent various KLGs. KLG1 is split into ‘KLG1progress’ (progressed to KLG2 in 96 months) and ‘KLG1Stable’. **(D)** Heatmap to show group mean differences in (C). We use *, ** and *** to denote significance levels *0.05, 0.01* and *0.001*, respectively, after Bonferroni correction for *28* knee regions in (B-D). Significance levels in (C-D) are shown based on Tukey adjusted pvalues.

In this analysis, we observed that cartilage thickness in all ROIs decreases significantly with age, with males typically possessing larger baseline thickness in most ROIs (except right posterior medial tibial region). While an increase in BMI negatively affected cartilage thickness in a majority of knee regions in males, the negative influence in females was restricted to the left and right posterior tibial and posterior lateral femoral regions, and right anterior and central medial femoral regions. Notably, all regions affected by BMI demonstrated a consistent negative age-BMI interaction, suggesting an increased negative effect of BMI on cartilage thickness as individuals age. The cartilage thickness reduction affected by both age and BMI was more evident in men compared to women among rOA-free participants. Regarding other factors, race typically negatively affected all ROIs with the exception of the anterior femoral region, and height typically exhibits positive effects, except for the anterior medial femoral, posterior lateral tibial, and left anterior medial tibial regions. In addition, femoral regions generally exhibited greater cartilage thickness than tibial ones.

Our results generally demonstrated bilateral symmetry between left and right knees, and confirmed a negative association between age and knee cartilage thickness at all sites (88). Previous research yielded inconsistent conclusions regarding age-sex interactions and cartilage loss. Cai, et al. (2019) (89) reported increased age-related lateral tibial cartilage loss in females, while Si, et al. (2021) (90) observed faster thinning in the lateral anterior and medial anterior femur in males. Our findings aligned more with Si, et al. (2021), likely due to our focus on rOA-free participants, in contrast to the higher proportion of OA-symptomatic participants in Cai, et al. (2019). Our data supported the notion of larger cartilage thickness in males (90-92). This might explain the lower OA prevalence rate in males (93), as thicker cartilage might offer better protection against shear stress (94).

For the effects of BMI, though numerous studies have examined the impact of BMI on cartilage loss in both sexes among OA patients (89), findings on the effects of BMI on cartilage thickness in healthy individuals are inconsistent. Some studies suggested an association between increased BMI and thicker cartilage (95, 96), while Keng, et al. (2017) (97) reported the opposite. Our investigation unveiled region-specific effects of BMI on cartilage thickness in rOA-free participants of both sexes. Moreover, the interaction between age and BMI remains largely unexplored. We found that age and BMI interact negatively in all femoral and tibial regions adversely affected by BMI. This might indicate an amplified detrimental impact of BMI on cartilage thickness with age. Such an effect could result from cumulative joint damage, increased load from higher body weight, as well as age-related metabolic and inflammatory changes. Additionally, we observed thicker knee cartilage among white and taller individuals, consistent with Connolly, et al. (2008) (98).

### Reference cartilage thickness charts highlight rOA progression patterns

We examined cartilage thickness differences among six KLG groups of knees, adjusting for confounding covariates. These groups included KLG2-4, KLG0, and KLG1. The KLG1 was further divided based on progression status to KLG > 1 as “KLG1progress” and “KLG1Stable”. The boxplots in **Figure 2C** and heatmap in **Figure 2D** display these differences across 28 regions. For the transition from KLG0 to KLG1, thickness increased in the posterior femoral (ROIs 1 and 6) and posterior medial tibial (ROI10) regions. Compared to the KLG1 stable group, the KLG1progress group showed additional increases in the posterior femoral, central lateral femoral (ROI2), and anterior medial tibial (ROI12) regions. In KLG1-2, we noted reductions in the posterior and central tibial (ROIs 7 and 8) and the central medial tibial and femoral regions. However, there was an increase in the left posterior lateral femoral thickness (ROI 1). For KLG2-4, cartilage thinning was observed in all ROIs except the left and right anterior femoral (ROIs 3 and 4) and left lateral femoral (ROIs 1 and 2). The most significant thinning occurred in the central medial tibial and femoral regions (ROIs 11 and 5). Besides, there is an increase in anterior lateral femoral region (ROI 3) from KLG2 to KLG3.

In summary, one of the earliest indications of progression to rOA was an increased thickness in the posterior femoral and posterior medial tibial regions, when transitioning from KLG = 0 to 1. As progression continued from KLG = 1 to 2, cartilage thinning occurs in posterior and central lateral tibial and central medial tibial regions with possible cartilage increase in the posterior lateral femoral region. During the transition from the KLG = 2 to 4, a significant decline in cartilage thickness across the majority of femoral and tibial regions was observed, while at the earlier transition (KLG = 2 to 3), cartilage thickening may occur in the anterior lateral femoral region. This progression trajectory outlined a population-level course of rOA progression.

In previous literature, OA pathogenesis may not strictly follow a linear path. Instead, a more complex picture, suggesting a bi-directional cartilage change during OA’s early stages, has been proposed (34). Cotofana, et al. (2012) (43) reported knees with early rOA displayed thicker cartilage specifically in the external femoral subregions of compartments with marginal osteophytes; Calvo, et al. (2001) (36) reported a progressive articular cartilage thickness increase in the femur at weeks 4, 6 and 8 but only at week 6 at the tibial region; findings from Hellio Le Graverland, et al. (2009) (44) suggested that, on average, female participants with KLG2 and KLG3 showed thicker and thinner cartilage than healthy controls, respectively, corroborating our observed thickness rise from KLG1 to KLG2 and a subsequent decline from KLG2 to KLG3. However, few studies have systematically examined detailed regional thickness changes across different phases and regions based on larger cohorts.

While our study identified several distinct patterns among different rOA categories, their predictive power needed to be examined. In the subsequent sections, we further explored the predictability of these patterns, including predicting rOA classification, incidence rOA and the initial-stage rOA transition.

### Reference cartilage thickness charts facilitate early rOA classification

Using 22 machine learning methods (more details in *Method* Section), we classified the knees in early-rOA phases (KLG ≤ 2) into three stages, including KLG ≤ 1, and KLG2 with and without progression into KLG3. This was carried out through two binary classifications, including classifying KLG ≤ 1 versus KLG = 2, and classifying KLG2 with versus without progression, whose performances were shown in **Figure 3** and **Figure 4**, respectively. The comparison of the area under the receiver operating characteristic (AUC), macro-F1 score and prediction accuracy (ACC) among any subset of demographic factors, raw regional cartilage thicknesses, and centile features depicted in **Figures 3B** and **4B**, and **Supplementary Tables S3-S5** and **S6-S8** using the combined demographic factors, raw regional cartilage thicknesses and centile features (refer to **Supplementary Table S2** for feature details). In both binary classifications, the combined set of predictors of demographic factors, raw regional cartilage thicknesses and centile features was optimal, which outperformed those reduced models without centile features, in terms of increased AUC, F1 score and ACC. Additionally, we conducted the classification to distinguish between (KLG ≤ 1) and (KLG ≥ 2) to evaluate our performance against existing literature. Results were shown in **Supplementary Figure S3 and Supplementary Tables S9-S11**.

**Fig. 3.**
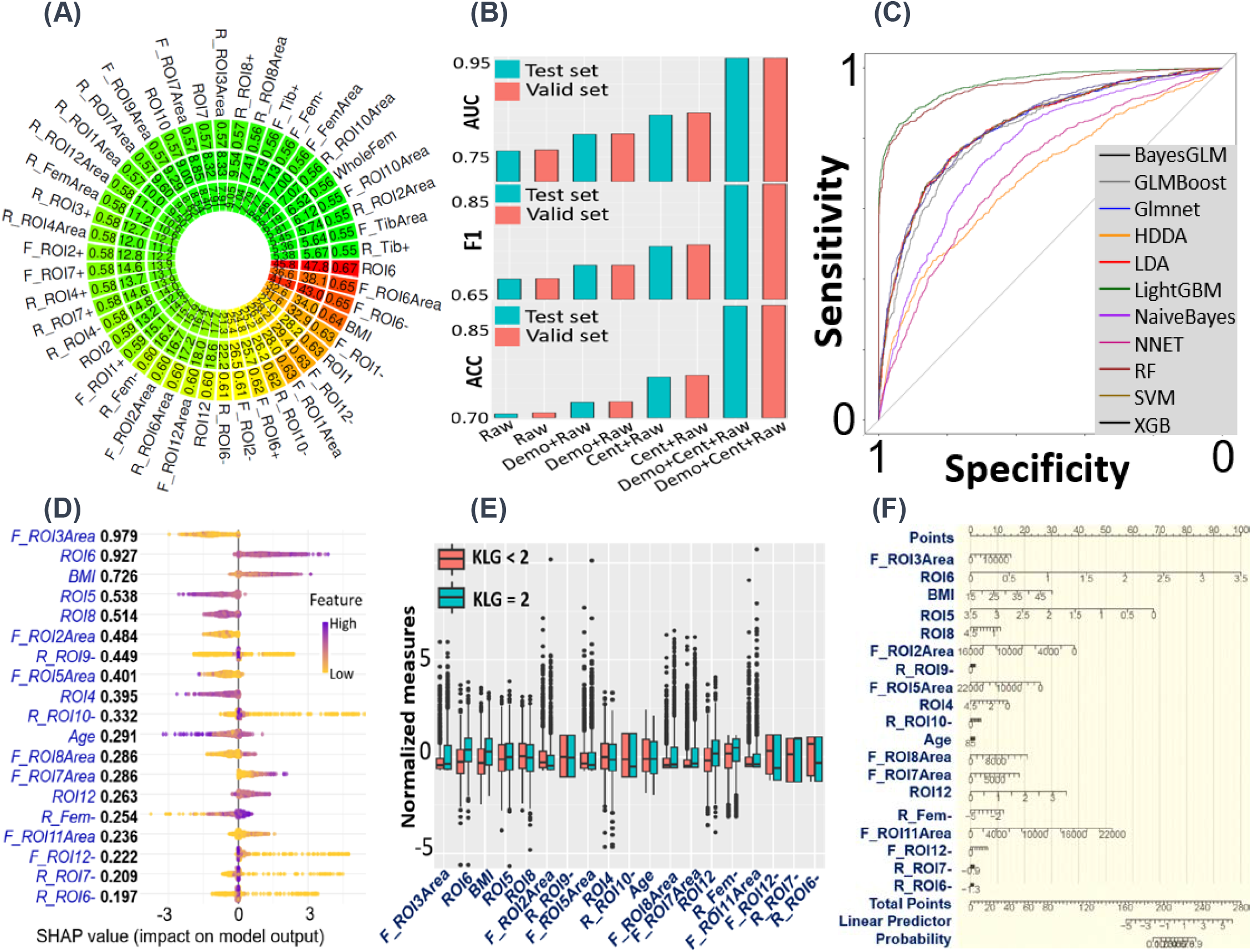
Results of rOA classification between KLG = 2 and KLG < 2. **(A)** Circular plot showing associations between radiographic OA (rOA) status KLG =2 and top predictors. Outer, middle, and inner circles signify Area Under the Receiver Operating Characteristic Curve (AUC), -Log10*p*, and - Log10(Bonferroni-adjusted *p*), respectively. Only results with AUC *> 0.55* are shown. **(B)** Barplots illustrating the median performance metrics (AUC, F1 score, Accuracy) across *22* machine learning methods using different predictor combinations (“Demo” for demographic factors, “Raw” for raw regional cartilage thicknesses and “Cent” for centile features). Performance for test (blue) and validation (red) datasets is indicated. **(C)** Receiver Operating Characteristic (ROC) curves of *11* models (each using random forest (RF) for variable selection) on the test set. The LightGBM model achieves the highest AUC and F1. **(D)** SHapley Additive exPlanations (SHAP) plot revealing impacts of top contributing 19 predictors (larger than 80% contribution in total) on the test set based on the LightGBM model. Predictors are sorted by their importance; for each predictor, each dot represents a patient’s SHAP value plotted horizontally, colored by the value of the feature from low (yellow) to high (purple). If purple and yellow dots are plotted at the lower and higher side, respectively, the KLG = 2 risk becomes higher as the feature increases. **(E)** Boxplots of the top predictors (normalized to mean zero and unit variance) for KLG = 2 (red) and KLG < 2 (blue). **(F)** Nomogram (100) depicting coefficients of (not standardized) top predictors in a joint logistic model. The x-axis (“points”) refers to the scores assigned to predictors based on their influence or weight in the predictive model.

**Fig. 4.**
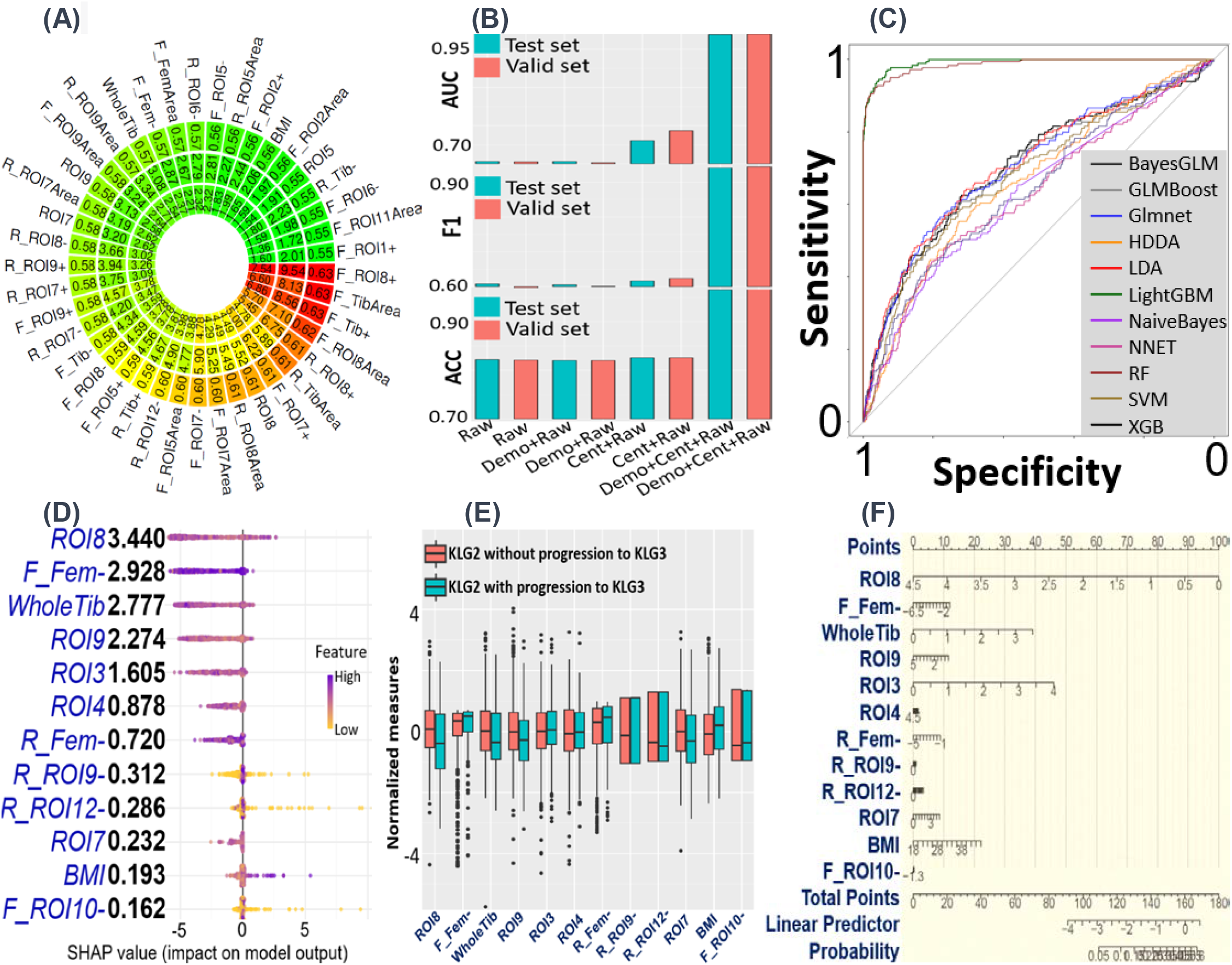
Results of KLG2-KLG3 progression prediction. **(A)** Circular plot showing associations between radiographic progression status and top predictors. Outer, middle, and inner circles signify Area Under the Receiver Operating Characteristic Curve (AUC), -Log10*p*, and -Log10(Bonferroni-adjusted *p*), respectively. Only results with AUC *> 0.55* are shown. **(B)** Barplots illustrating the median performance metrics (AUC, F1 score, Accuracy) across *22* machine learning methods using different predictor combinations (“Demo” for demographic factors, “Raw” for raw regional cartilage thicknesses and “Cent” for centile features). Performance for test (blue) and validation (red) datasets is indicated. **(C)** Receiver Operating Characteristic (ROC) curves of the *11* models (each using random forest (RF) for variable selection) on the test set. The LightGBM and RF models achieve the highest AUC and F1, respectively. **(D)** SHapley Additive exPlanations (SHAP) plot revealing impacts of top 12 predictors (> 90% contribution) on the test set based on the RF model. Predictors are sorted by their importance; for each predictor, each dot represents a patient’s SHAP value plotted horizontally, colored by the value of the feature from low (yellow) to high (purple). If purple and yellow dots are plotted at the lower and higher side, respectively, the progression risk becomes higher as the feature increases. **(E)** Boxplots of the top predictors (normalized to mean zero and unit variance) for progressive KLG2 (red) and stable KLG2 (blue). **(F)** Nomogram (100) depicting coefficients of top predictors in a joint logistic model. The x-axis (“points”) refers to the scores assigned to predictors based on their influence or weight in the predictive model.

For the classification of KLG ≤ 1 against KLG = 2, the LightGBM method, with random forest (RF) permutation importance feature selection, proved most effective (**Figure 3C**), achieving both the highest AUC of *0.953* (*σ = 0.001)* and F1 of *0.887* (*σ = 0.003*). Various plots, including the association circular plot (**Figure 3A**), SHapley Additive exPlanations (SHAP) plot (**Figure 3D**), pairwise comparison boxplots (**Figure 3E**), and nomogram (**Figure 3F**), elucidated the predictors’ significance, effect sizes, prediction contributions, and joint effects. In particular, cartilage thickening in the posterior medial (ROI 6) and anterior lateral (ROI 3) femoral regions, cartilage thinning in the posterior and central lateral tibial (ROIs 7 and 8) and central medial femoral (ROI 5) regions, coupled with an increased BMI, contributed to the classification of KLG = 2.

In differentiating KLG2 based on progression, the RF and LightGBM (with random forest feature selection) achieved the highest F1 score and AUC, respectively (**Figure 4C**). In particular, the RF achieved an AUC of *0.979* (*σ = 0.002)* and F1 score of *0.941* (*σ = 0.006*). The corresponding circular plot, SHAP plot, boxplots, and nomograms were displayed in **Figure 4A, 4D, 4E** and **4F**, respectively. A decrease in cartilage thickness in the lateral tibial regions (ROIs 7, 8 and 9), an increase in the anterior femoral regions (ROIs 3 and 4), and a heightened BMI increased the risk of rOA progressing to KLG3.

When distinguishing between KLG ≤ 1 and KLG ≥ 2, the LightGBM method, combined with GLMBoost importance-based feature selection, yielded the best performance with an F1 score of 0.818 (*σ* = 0.001) and an AUC of 0.900 (*σ* = 0.003) (**Supplementary Figure S3C**). Factors contributing to a classification of KLG ≥ 2 include cartilage thickening in posterior femoral (ROIs 6 and 1) region, thinning in central tibial (ROIs 11 and 8), central medial femoral (ROI 5) and the overall femoral and tibial regions, increased BMI and age, and identifying as female, and black (**Supplementary Figure S3A, S3D, S3E** and **S3F**).

In existing literature, there were studies (53) on the auto-classification of 5 KLG scores. However, few have specifically focused on differentiating between KLG ≤ 1 and KLG = 2 or between KLG2 with and without progression. In addition, our performance in classifying KLG ≤ 1 against KLG ≥ 2 was comparable to the 3D CNN model proposed by Guida et al. (2021) (99) which achieved an F1-score of 0.831 and an AUC of 0.911. Our approach, however, offered enhanced simplicity and interpretability, and was validated on a larger cohort.

### Reference cartilage thickness charts predict the incidence and initial-stage rOA

In this section, we distinguished between incidence rOA (KLG increasing from < *2* to ≥ *2*) and initial-stage rOA (KLG from 0 to 1) transition. We evaluated the performance of 22 machine learning models, using a combination of demographic factors, raw regional cartilage thicknesses, and centile features. For the incident rOA prediction, the optimal models (hightest F1 score) incorporated all these predictor variables, while for initial-stage predictions, the optimal model incorporated predictor variables without demographic variables. In both cases, they outperformed reduced models that did not include centile features. Detailed model performances for incidence and initial-stage rOA predictions can be found in **Supplementary Figures S4B** and **S5B**, and **Supplementary Tables S12-S14** and **S15-S17**. While ROC curves of the top models were similar (**Supplementary Figure 4C**) for incident rOA prediction, there were differences for initial-stage prediction (**Supplementary Figure 5C**). The linear discrimant analysis (LDA) model, coupled with Glmboost for feature pre-screening (AUC *= 0.813, σ = 0.002* ; F1 *= 0.685, σ = 0.004*), and the logistic regression (Glmnet) method, coupled with random forest feature pre-screening (AUC *= 0.653, σ = 0.021*; F1 *= 0.639, σ = 0.036*) were optimal models for the incidence and initial-stage rOA transition prediction, respectively. The marginal associations including AUC, p-values and Bonferroni-corrected p-values of top predictors (AUC *> 0.7*) with rOA are illustrated in **Supplementary Figures S4A** and **S5A**, the importance of top predictors on the test set are shown in the SHAP plots (**Supplementary Figures S4D** and **S5D)**, and boxplots of group differences of normalized (zero mean and unit variance) top predictors were shown in **Supplementary Figures S4E** and **S5E**. These predictors’ joint effects were shown in nomograms (**Supplementary Figures S4F** and **S5F**). From those results, we observed that the larger BMI, along with increased cartilage thickness at the posterior and central lateral femoral regions and reduced cartilage thickness at the overall tibial and femoral regions served as top contributors of rOA early-stage progression, while increased cartilage thickness at posterior lateral, posterior medial femoral regions, and reduced cartilage thickness in the central medial femoral region were initial-stage rOA progression indicators.

These findings were in line with previous research demonstrating the relationship between BMI and OA development (101, 102). The swelling of lateral femoral cartilage thickness in predicting early-stage rOA progression paralleled findings from studies such as Erhart, et al. (2008) (103). Furthermore, our study revealed the increase in the posterior medial and the reduction in the central medial femoral regions as potential biomarkers of intial rOA. This underlined the importance of further studies to explore underlying mechanisms.

### Reference cartilage thickness charts aid in local disease mapping for rOA progression

We analyzed the difference of transformed local centile scores representing cartilage thinning and thickening (details in *Method* Section) between participants with and without rOA progression, delineated across five KLG groups in the femoral and tibial regions (**Figure 5A**). **Figure 5B** illustrates differences and significance of comparisons shown in **Figure 5A**, indicating locations where cartilage thickness differed between participants with and without rOA progression at each radiographic stage. In particular, we observed increased cartilage thickness in the posterior medial and posterior lateral femoral regions and a reduction in the central medial femoral region in the dashed black boxes in **Figure 5A**. Furthermore, we discerned (**Figure 5B**) a substantial reduction in the lateral tibial, increase in posteror medial femoral regions and minor reduction in the anterior femoral regions in participants with rOA progression at KLG *= 2*, which could potentially signify a progression towards KLG = 3. Similarly, a larger reduction (**Figure 5B**) in the medial tibial region among KLG3 participants might suggest further progression into KLG = 4. We prepared a movie available at https://tli3.github.io/KneeOA/ which showcased dynamic changes in local centile scores for four subtypes of rOA progression during all visits: KLG = 0 → KLG = 1; KLG = 0 → KLG = 1 → KLG = 2; KLG = 0 → KLG = 1→ KLG = 2 → KLG = 3; KLG = 0 → KLG = 1 → KLG = 2 → KLG = 3 → KLG = 4. This demonstrated how local centile scores of the four rOA progression patterns changes across time. While there is extensive literature on the topic of rOA progression prediction, few studies specifically address an overall local progression of rOA.

**Fig. 5.**
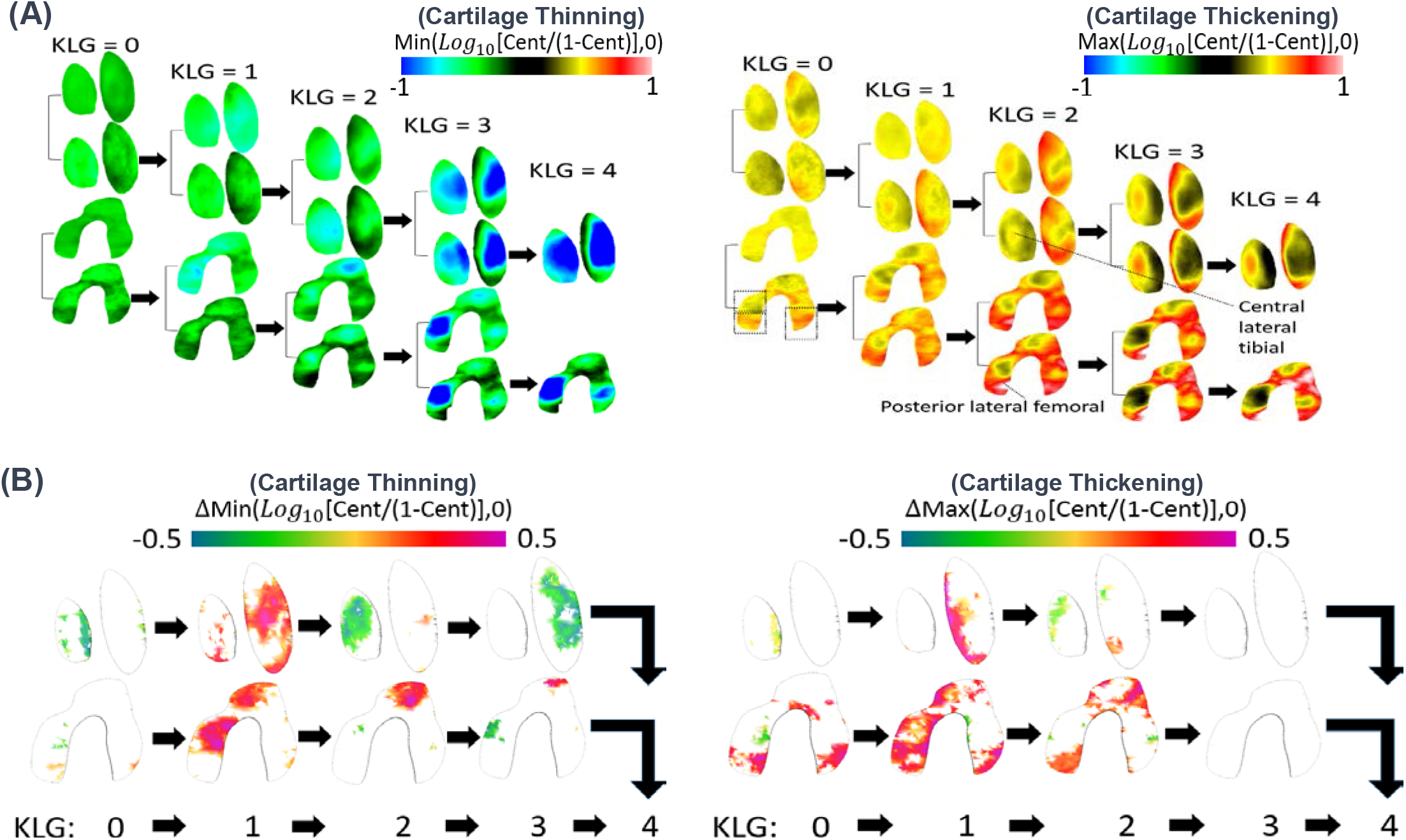
Visualization of the local centile score map for different KLG progression patterns. **(A)** A comparison of mean local centile scores, which were logit10-transformed (*log10(x/(1-x))*), between participants with and without rOA progression across five KLG groups in the femoral and tibial regions. These scores were separated into positive and negative components, representing bidirectional thickness changes (reduction and increase). **(B)** Differences of each comparion in panel (A), showing locations with FDR-adjusted *p < 0.2*. ΔMax(*Log10*[Cent/(1-Cent)],0) = Max(*Log10*[Cent2/(1-Cent2)],0)-Max(*Log10*[Cent1/(1-Cent1)],0), where Cent2 and Cent1 are local centile scores with and without progression, respectively (ΔMin defined similarly). A pink (or green) color in cartilage thinning indicates a decrease (or increase) in OA-induced cartilage reduction compared to non-progression; a pink (or green) color in cartilage thickening indicates an increase (or decrease) in OA-induced cartilage thickening compared to non-progression.

## Discussion

### An integrative toolset for rOA predictive analyses

Traditional knee rOA association and prediction analyses (30-33) have primarily relied on manual readings of MR images, which often limit the study sample size and statistical power, potentially leading to conflicting results. Modern deep-learning models (45, 46), while offering accurate rOA prediction, pose challenges for clinical interpretability. In response to these challenges, we have developed an integrative toolset that combines the benefits of both traditional and modern approaches. This toolset includes DL-based LCT maps extracted from knee MR DESS image data, rOA-free nonlinear growth charts of knee cartilage thickness, enhanced statistical methods for extracting centile features and the implementation of ML-predictive modeling. These components work in harmony to allow more comprehensive OA diagnosis and prognosis. All codes are written in R/4.2.2 which we will make publicly accessible at www.github.com. To further supplement our toolset, we have proposed a variety of visualization methods, including circular plots, SHAP plots, boxplots, and nomogram plots. These are integrated with local diagnosis visualizations, providing a means to capture and visualize regional abnormalities during disease progression. Compared to previous research which has often examined cartilage thickness changes during rOA progression in a broader scope, our developed toolset advances precision by identifying localized alterations in cartilage thickness among participants at various radiographic stages with or without future progression. This enhanced our prior work (64), which although successfully detected local dynamic abnormal regions and rOA subclassifications, did not predict future rOA progression.

On the other hand, the effective contribution of our methodology is reflected in its robust performance and high prediction accuracy, with AUC values of 0.953, 0.900, 0.979 and 0.813 and F1 scores of 0.887, 0.818, 0.941, and 0.685, for rOA classification of KLG ≤ 1 versus KLG = 2 and KLG ≤ 1 versus KLG ≥ 2, and prediction of KLG2-3 transition and incidence rOA transition, respectively. We here include the macro-F1 score into our evaluation metrics, which effectively balances precision and recall, providing a more conservative evaluation in scenarios with imbalanced data (104). Despite being frequently used in prior studies (48, 49, 52, 55, 60, 63), metrics like accuracy and AUC can be sensitive to the class distribution and may result in overly-optimistic results in imbalanced scenarios, as the model may perform well on the majority class but poorly on the minority class. In previous literature, Thomas, et al. (2020) (53) and Guida et al. (2021) (99) conducted the same classification task as ours in classifying KLG ≤ 1 against KLG ≥ 2, whose reported mean F1-score were 0.80 and 0.831 based on deep learning methods (our result was 0.818). Tiulpin, et al. (2020) (57) reported a macro-F1 of less than 0.60 and 0.57 for rOA multi-class classification. In terms of predicting OA progression, Widera, et al. (2020) (59) reported a weighted F1 of 0.69 based on the OAI data, while Chan, et al. (2021) (61) achieved F1 scores of 0.837 and 0.810 for predicting OA incidence and deterioration, respectively, indicating higher prediction accuracy than our model. However, it is worth noting that their model used additional symptomatic data, history of knee injury and surgery, metabolic syndrome and living habits, for prediction of OA progression characterized by JSW and WOMAC pain score, while our work focused on easily-measurable factors such as age, sex, BMI, race, and height to predict rOA progression.

In addition, previous literature on early prediction of OA generally focuses on the definite-OA phase. Studies predicting the rOA initial transition remain less explored. Shamir, et al. (2009) (105) reported the automatic detection of doubtful rOA (KLG = 1) with a relatively low accuracy of *57%*; our analyses predicted the transition to KLG = 1 from KLG = 0 with a higher accuracy of *65.9% (σ = 0.027)*. Predicting initial-phase rOA onset would provide insights of the underlying mechanisms of early OA. Furthermore, in our study, the consistency of the identifed regions and signals observed across diverse analyses, including regional association analyses, future predictive analyses, and local disease region analyses lends robustness to our findings.

### Insights and findings from a clinical perspective

In knee OA patients, prevalent symptoms of pain and joint stiffness often originate from innervated tissues such as the synovium and bone (106, 107), instead of the typically avascular and aneural articular cartilage. Yet, cartilage degradation is closely linked to synovitis, bone marrow lesions, and osteophyte formation. The degradation of cartilage can release inflammatory mediators that trigger synovitis, which in turn can accelerate cartilage degradation, thus perpetuating a destructive cycle (108, 109). Moreover, as cartilage degrades, the resulting increased load and altered joint mechanics can lead to bone marrow lesions, detectable through imaging. As this process unfolds, cartilage thinning often occurs as one of the early detectable signs of OA. Thus, assessing changes in cartilage thickness using MRI serves as a valuable tool for diagnosing and tracking OA progression.

While considerable research has delineated the association between BMI, age, and cartilage thickness (89, 90, 95-97), studies on the age-BMI interplay remain scarce. We construct an age-BMI growth chart for knee cartilage thickness among rOA-free participants, incorporating complex age-BMI interactions. The observed interactions between age and BMI in our study support the notion that high BMI values may accellerate aging for rOA-free knee cartilage.

Previous literature (34, 40-44) has observed that cartilage thickening may act as important indicators in early OA. For instance, it was observed (34) that the frequency of knees classified with medial femorotibial cartilage thickening was equal to that of medial thinning in knees with KLG = 2, whereas cartilage thinning was 2.6 times more frequent than thickening in knees with KLG = 3. Despite these observations, understanding of cartilage thickening in early stages of OA is limited, necessitating further validation through larger-scale studies. Commonly used OA imaging scores like MOAKS and WORMS primarily focus on cartilage loss, which may lead to underrepresentation of cartilage thickening in early stages. Furthermore, the potential of cartilage thickening to serve as a predictive biomarker for rOA progression remains unclear. Our results provide evidence to suggest that cartilage thickening may occur at the initial and earlier radiographic grades and may present a biomarker for rOA progression.

In summary, our study has shed light on the potential of combined statistical-ML approaches to provide valuable insights into rOA progression and its early indicators. Further studies are warranted to confirm our findings and explore the clinical implications for early detection and prognosis of rOA.

### Limitations and future work

Certain limitations exist within our study that should be highlighted. Within our scope of study, our predictor variables were focused on easily-measurable factors such as age, sex, BMI, race, and height, which can be obtained without additional clinical examinations. Nevertheless, other influential factors, such as knee injury, knee alignment, laxity, quadricep muscle strength, physical activity, genetic predisposition, and overall health conditions, could be integrated into future models to enhance disease incidence and progression predictions. Second, while we have chosen to predict rOA progression, a broader scope could include the prediction of JSN, symptoms, cartilage lesion incidence, and TKR. As rOA only forms a component of OA, pain scores could also be considered for the OA prediction (29) instead of rOA. Third, we have constructed cartilage thickness growth charts for rOA-free participants in the OAI, which are not the same as healthy controls. There lies potential for future research to build these growth charts for healthy control participants without knee pain, which could be beneficial for comparative studies. Finally, while our models utilize scan features from one single scan to predict future rOA progression, exploring advanced statistical models that incorporate features from all historical scans, like autoregressive models, could offer deeper insights.

## Method

### Study population

The Osteoarthritis Initiative (OAI; https://nda.nih.gov/oai/) is a prospective, multi-center, longitudinal study initiated in 2004. It recruited 4,796 participants aged 45 to 79 years, who were either at high risk of or already had symptomatic knee OA (82). Participants were followed for up to 96 months, with imaging and clinic visits scheduled at baseline, 12 months, 24 months, 36 months, 48 months, 72 months and 96 months (110). We have observed significant difference in knee cartilage thickness between KLG = 0 and KLG = 1 participants, supporting the use of rOA-free participants for the creation of reference group charts. Generally, we used the 957 rOA-free participants as the training data for the construction of the rOA-free reference cartilage thickness growth charts. Participants included in all the other participants were used to test the prediction performance of rOA progression based on the reference charts. Given the potential bilateral symmetry in rOA progression (111), it is not recommended to consider a knee with KLG0 as rOA normal if its contralateral knee exhibits a KLG > 1. Based on this, we excluded participants if they met any of the following criteria: (i) one knee consistently presents with KLG ≤ 1 across all visits while the contralateral knee has KLG > 1 at any visit, (ii) one knee consistently shows KLG = 0 across all visits while the contralateral knee displays KLG = 1 at any visit, (iii) knee MRIs are missing or are of poor quality across all visits, or (iv) participants of minority races, those identifying as hispanic, or those not identifying as either white or black. Sample size and other demographic information for participants at their baseline visits in different subpopulations were summarized in **Table 1** (based on the KLG of right knees).

### Magnetic Resonance Images and preprocessing

Longitudinal assessments and measurements were performed on 4,796 participants at up to seven time points by using MR imaging data. The imaging data was collected using four 3.0 Tesla Siemens Trio MRI scanners over a four-year follow-up period. The use of 3T-MRI scanners and standardized protocols, as opposed to 1.5T-MRI, offered greater signal-to-noise and contrast-to-noise ratios due to its larger field strength. The OAI applied various imaging acquisition protocols. We specifically used those MRIs employing the sagittal 3D Dual Echo Steady State (DESS) technique with selective water excitation (WE), which had been demonstrated superior for cartilage discrimination (110). Although there were a few exceptions, the majority of these images had dimensions of 384 × 384 x 160 and a voxel resolution of 0.36 × 0.36 × 0.7 mm^3^. In prior research, we developed the Dynamic Abnormality Detection and Progression (DADP) pipeline (64) and an open-source knee-OAI analysis toolkit https://github.com/uncbiag/OAI_analysis_2 for the creation and analysis of knee thickness maps. In this study, the OAI MR images were preprocessed (**Figure 1A**) through the image analysis module including the following four steps: segmentation and meshing; 3D thickness map computation; registration; and thickness map projection. The output of this step was a two-dimensional (2D) thickness map in a common 2D atlas coordinate system for each OA MR image.

### KLG measures for radiographic severity

The KLG system is a scoring system used to evaluate the severity of knee OA on radiographic images. The scale ranges from 0 to 4, with 0 indicating the absence of changes and 4 indicating the presence of substantial osteophytes, marked by joint space narrowing, severe sclerosis, and evident deformity of the bone contour. The KLG scores were obtained from radiographs taken in the standing postero-anterior position using the fixed-flexion protocol. This protocol involves acquiring bilateral X-rays while the patient stands with knees flexed between 20-30 degrees and feet internally rotated 10 degrees. To ensure consistent patient positioning, the degree of knee flexion and foot rotation was fixed using a plexiglass positioning frame (SynaFlexer, Synarc, Inc, San Francisco, CA, USA) (112).

### Construction of the FKTR charts

After obtaining 2D maps of cartilage thickness for all individuals in the common coordinate system, we used statistical models to construct the rOA-Free Knee Thickness Reference (FKTR) charts. The charts aimed to characterize the aging trajectories of LCT maps using data from 957 rOA-free individuals. To do this, an experienced musculoskeletal radiologist (DN) manually annotated 12 ROIs with 6 for the femoral and 6 for the tibial cartilages on the standard 3D atlas space according to their physical locations. These ROIs were then mapped to the 2D atlas space (**Figure 1B**). Next, we fitted both regional-average and local cartilage thickness regression models, taking into account the demographic covariates including age, sex, BMI, height, and race of the individuals as covariates. Note that in this step, clinical covariates were considered contributing factors rather than confounding factors. We used a Generalized Additive Mixed Model (GAMM) (113), which allowed us to model the nonlinear and dynamic nature of cartilage thickness development during aging. Let *y*_*i,v*_(*t, b*) denote the cartilage thickness for the *i*-th subject at location *v* and age *t*, and with BMI *b*, with *i* = 1, …, *n*. Specifically, the GAMM model can be formulated as the model *M*1:

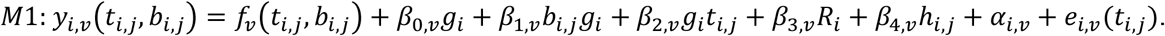

Here *g*_*i*_ and *r*_*i*_ represent the sex and race for subject *i*, with *g*_*i*_ = 1 for male and 0 for female and *R*_*i*_ = 1 for black and 0 for white groups since most of the participants are white, *b*_*i,j*_ and *h*_*i,j*_ represent the BMI and height for the subject *i* at age *t*_*i,j*_, respectively, and *α*_*i,v*_ and *e*_*i,v*_ (*t*_*i,j*_) denote the random intercept effect and random Gaussian noise for subject *i* at location *v* and age *t. β*_0,*v*_, *β*_1,*v*_, *β*_2,*v*_, *β*_3,*v*_, and *β*_4,*v*_ are coefficients of the sex, BMI-sex and sex-age interactions, race and height, respectively. Our rationale for examining the complex interactions between age and BMI is that both serve as important continuous indicators of aging and knee OA. Taking the variables described above, the two-dimensional functions *f*_*v*_(*t, b*) were estimated by full tensor product smoothing using cubic splines (114) to model the nonlinear age-BMI interactions nonparametrically. Smoothing hyperparameters in *f*_*v*_ (*t, b*) were chosen by maximizing the marginal likelihood. ANOVA F-statistics were calculated to determine the significance of the main effects and BMI-sex, age-sex interactions. Two alternative models were compared with model M1 in the **Supplementary Text**. All the above estimations and 95% simultaneous confidence intervals were calculated through the mgcv package (115) with R version 4.2.2. This analysis was solely based on the rOA-free participants.

### A comparison of cartilage thickness progression between different rOA groups

Participants with KLG = 0 and KLG = 1 were often grouped together as rOA-free due to the subtle distinctions between these grades (116). In our study, we separated them to enable detailed comparisons across OA progression phases. First, we adjusted the cartilage thickness for all confounding factors, by removing all fixed effects of rOA-free participants from model M1. Afterwards we fit the following linear mixed model *R*1

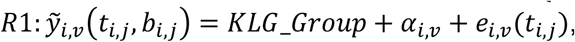

where 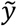 is the residualized cartilage thickness, and *KLG_Group* is a categorical variable which includes six KLG group, the KLG0, KLG2, KLG3, KLG4, “KLG1progress” (progression to KLG > 1) and “KLG1Stable”, and *α*_*i,v*_ and *e*_*i,v*_ (*t*_*i,j*_) are Gaussian random effects and random noise, respectively. One-way ANOVA F tests statistics were used to test the effect of KLG on the residualized cartilage thickness, accounting for mixed effect models via Satterthwaite’s degrees of freedom method (117). These tests were performed across 28 regions, including 12 ROIs and the overall femoral and tibial regions in both the left and right knees. P-values were adjusted for 28 independent tests using the Bonferroni correction. For regions with significant KLG group effect, posthoc analyses were performed to test pairwise differences using the Tukey test to correct for multiple comparion of pairs. This analysis was performed through the lme4 and emmeans packages with R version 4.2.2, and was based on all scans from rOA-prevalence and KLG0-1-stable participants.

### Local centile score extraction

In the preceding subsection, we constructed reference developmental trajectory charts to depict knee growth patterns in individuals without rOA. Using these charts as a baseline, we compared and evaluated the developmental trajectories of knee cartilage thicknesses from participants in different groups. We introduced the **Local centile Scores (LCS)** to assess the extent of local abnormalities/extremities for a given knee. Specifically, we defined the LCS at each location *v* as the centile of cartilage thickness among all rOA-free knees, adjusted for demographic confounding covariates. After accounting for these confounders, we ranked the residualized cartilage thickness of the knee relative to the residuals of all rOA-free knees at each *v*, and then convert these ranks to centiles. The LCS could be calculated in two ways, which account for fixed effects only from confounding covariates (Method <u>F</u>) or both the fixed and participant-level random effects (Method <u>R</u>), respectively. More details and formulations were shown in the **Supplementary Text**. Similar methods can be found in the centile scores of Bethlehem, et al. (2022) (118).

### Association analyses between rOA progression and LCS

First, the LCS underwent a logit transformation, a procedure that extended the values near the extremes of the centile (closer to 0 or 1),

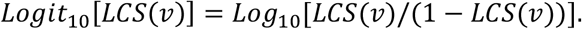

The median is naturally mapped to *Logit*_10_0.5 = 0. This transformation essentially increases the sensitivity of our model to extreme centiles, allowing us to assess the degree of departure from the median cartilage thickness in both directions. We then bifurcated these transformed scores into negative and positive components. This segregation represents the bidirectional changes in cartilage thickness: the **cartilage thinning LCS** and **thickening LCS**,

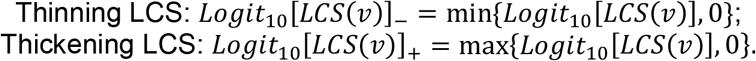

Specifically, the positive components reflect instances where cartilage thickness is greater than the median, while the negative components capture instances where cartilage thickness falls below the median. Next, we tested the difference of thinning and thickening LCS between KLG0 knees with and without KLG1 progression, between KLG1 knees with and without KLG2 progression, between KLG2 knees with and without KLG3 progression, and between KLG3 knees with and without KLG4 progression, using two-sided two-sample t-tests. P-values were FDR-corrected for all pixels *v*.

### Centile feature extraction

We proposed a novel concept of the **global centile scores (GCS)** to assess the global abnormality/extremity of a given knee across all locations. Specifically, the GCS is defined as the mean centile of **the LCS spatial distribution** among rOA-free knees, where the LCS spatial distribution is characterized by the cumulative distribution function (CDF). Denote the CDF of *LCS*(*v*) for *v* ∈ *V* as *F*(*·*), which can be formulated as

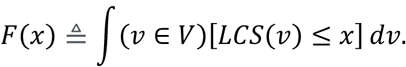

The centile function *GCS* (*x*) (in region *V*) was defined as the centile of *F*(*x*) among that of all rOA-free knees,

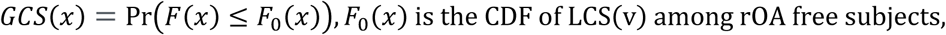

and the final GCS is defined as the mean of *LCS*(*x*) function (mean centile function),

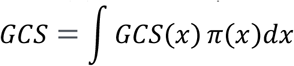

where *π*(*x)* was a Bayesian prior on the interval [0,1], such as a uniform distribution on [0,1], which is used to specify which percentile of the LCS distribution is more important for global diagnosis.

This method was motivated by the cluster-extent based thresholding method (119-121). The CDF can also be interpreted as the area where LCS is less than the threshold *x*. For example, when *π*(*x*) = 1 at *x* = 0.05 and 0 otherwise, the CDF is the “disease region” size determined by the “p-value” < 0.05. In this sense, the GCS is an extension to cluster-extent p-value-based inference, with a fix significance level. In our setting, *π*(*·*) was data-driven (**Supplementary Text**). Similar to the LCS, we calculated the **cartilage thinning GCS** and **thickening GCS** by

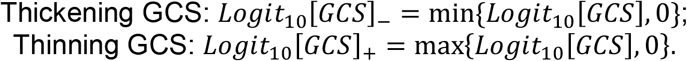

Besides the GCS, we extracted other centile features, the integrated CDF (ICDF), based on the CDF of the LCS. The ICDF was defined as

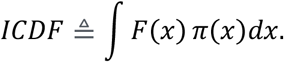

This integration reflects the mean cartilage thinning area across all threshold *x*. Both a larger ICDF and GCS imply a reduced cartilage thickness. The thickening GCS, thinning GCS and ICDF features were extracted across 14 regions (12 ROIs and the overall femoral and tibial regions). In total, for each knee we extracted 84 centile features, including 14 Thinning GCS, 14 Thickening GCS, and 14 ICDF features, with each based on the LCS calculated from Method <u>F</u> or Method <u>R</u> (3 × 2 × 14 = 84). Detailed feature intepretations were given in the **Supplementary Table S2**.

### Classification and prediction tasks

Given the extensive analyses undertaken in this work, we have chosen to focus on the right knees for the subsequent classification and prediction tasks. Nevertheless, the established predictive framework is equally applicable to the left knees.

In the first classification, we categorized knees in the early-rOA phases (KLG ≤ 2) into three distinct stages: KLG ≤1, KLG2 progressing to KLG3 in 48 months, and KLG2 without such progression. For KLG ≤1 (referred to as Class 1-1), we chose the knee from 48 months prior to the last visit in KLG0-1-stable participants, ensuring the KLG remained ≤ 1 in the subsequent 48 months. This resulted in 1,362 knees from 1,362 participants. For KLG2 without progression (Class 1-2), we selected the knee from 48 months prior to the last visit in KLG2-stable participants, yielding 804 knees from 804 participants. For participants undergoing a KLG2-3 transition (Class 1-3), we chose the knee from the final visit before its transition to KLG3, yielding 179 knees from 179 participants.

In the second classification, we aimed to distinguish between knees with KLG ≤1 (Class 2-1) and those with KLG ≥ 2 (Class 2-2). This task is a typical rOA classification problem, which was performed to validate our methods against existing literature. In particular, we incorporated all 449 KLG4 knees, and randomly selected 450 knees each from KLG0, KLG1, KLG2, and KLG3 categories to balance the sample size. For each participant, only one knee was randomly chosen, with the exception of those in the KLG4 category due to limited knees with KLG4. This configuration aligns well with the settings in Guida et al. (2021) (99).

In the rOA-incidence prediction, we predicted the transition into KLG2 in 48 months based on its current knee with KLG < 2. For knees with no such transition (Class 3-1), we selected the knee from 48 months prior to the last visit in KLG0-1-stable participants. This resulted in 1,362 knees from 1,362 participants. For knees with transition (Class 3-2) we chose the knee from the final visit before its transition to KLG2, accounting for 213 knees from 213 participants.

In our rOA initial transition prediction, we aimed to predict the transition to KLG1 within 48 months for knees currently at KLG = 0. For knees with no transition (Class 4-1), we selected the knees from 48 months prior to the last visit in KLG0-1-stable participants, yielding 957 knees from 957 individuals. On the other hand, for knees with transition (Class 4-2), we used data from the last scan before their transition to KLG1, resulting in 46 knees from 46 participants. Given the significant sample size imbalance between the two classes, we randomly downsized the Class 4-1 group, by randomly selecting 46 knees for a balanced 46:46 classification task.

### Predictive modeling and hyperparameter selection

For better interpretation, we broke down the above three-stage classification of KLG ≤ 2 into two binary classification problems: differentiating Class 1-1 from Classes 1-2 and 1-3, and distinguishing Class 1-2 from Class 1-3. In all the classification and prediction tasks, we extracted global centile features within each of the 14 ROIs, which were combined with the 14 raw mean cartialge thickness, age, sex, BMI, height, and race, which summed up to 103 potential biomarkers (**Supplementary Table S2**). The KLG of current knee was used as an additional predictor only in the rOA-incidence prediction. For each prediction task, we employed a 10-fold nested cross-validation to randomly divided knees into training, validation, and test sets.

We adopted a standard two-step predictive modeling approach. Initially, we computed the random forest permutation importance (122) and the GLMBoost importance (123) of each feature using the training dataset. Subsequently, features were ranked based on one of these importance scores. Afterwards, top *S* features were selected, which were fed into 11 baseline models, including the elastic net (Glmnet) (124), Glmboost (125), single-hidden-layer neural networks (NNET) (126), Naive Bayes (NaiveBayes) (127), Bayesian Generalized Linear Model (BayesGLM) (128), random forest (RF) (129), XGBoost (XGBTree) (130), Kernel-based Support Vector Machine (SVM) (131), Linear Discriminant Analysis (LDA) (132), High-Dimensional Discriminant Analysis (HDDA) (133) and LightGBM (134). The optimal hyperparameter *S* was determined by maximizing the AUC on the validation dataset. All other hyperparameters, such as learning rate, maximum depth and regularization parameters, were optimized using an internal 10-fold cross validation within the training data. Pairing the two feature selection methods with the 11 baseline models yielded 22 distinct predictive approaches. The method achieving the highest F1-score on the validation data was adopted as our final model.

### Nested cross validation and model evaluation

To prevent overfitting, we used a three-layer-nested 10-fold cross-validation (CV) method to assess various predictive models. This method comprised three nested loops, all implementing a 10-fold cross-validation. Specifically, we partitioned all participants into 10 folds in the outer loop. Reserving one fold as an independent test set, the remaining folds were further divided into 10 subfolds in the middle loop. Choosing one subfold as the validation, the remaining folds were further divided into 10 internal subfolds in the inner loop. The outer loop was designed to give an unbiased performance evaluation of the model, the middle loop was used for determining the optimal number of selected features, and the inner loop was used for tuning all other hyperparameters. Because the test set was cycled through the 10 folds in the outer loop, each subject was evaluated once as an independent test subject. The prediction ACC, precision, recall, AUC, and F1-score for each machine learning model were reported. Furthermore, the three-layer-nested 10-fold CV was repeated five times to assess the standard deviation of these evaluation metrics.

### Visualizing results for enhanced clinical interpretation

Circular plots were shown to include the association results between each feature and the response (class label), which specifically, included the p-values based on two-sided two-sample t-test, FDR-adjusted p-values, and the univariate AUC. We then performed the prediction tasks based on various feature combinations, such as raw cartilage thickness features only, the combination of raw and centile features and the combination of raw, centile and demographic features. Barplots were displayed to compare the AUC, F1 and ACC of different combinations, based on their corresponding final incorporated models. ROC curves was used to compare the specificity and sensitivity among *22* predictive approaches.

The SHAP summary plot were used to visualize both the SHAP importance (135) score of each feature and the feature’s effect on the predictions. The y-axis represented the features sorted by their SHAP importance scores, while the x-axis reflects the Shapley value. Each dot on the plot represents a Shapley value for a specific feature and instance. An increase in the Shapley value suggests that, on average, including the feature would increase the predicted odds ratio for the instance. The color gradient indicates the feature’s value, varying from low to high. Boxplots were used to display the group differences for features with top SHAP importance (contributions *> 90%* or *80*%).

Finally, a nomogram chart was displayed to illustrate the combined effects of the top features, based on the assumption of a linear relationship between the log odds ratio and the features in a logistic regression model. In particular, the x-axis represent the points (100). Each predictor was assessed by moving vertically on the chart to find the corresponding points based on the patient’s score, yielding the individual score. The total score was determined by summing up all the individual scores. Then, one moved vertically down from the “Total Points” scale to ascertain the probability of being identified as cases. Although the linear relationship did not typically provide the optimal fit, this nomogram chart roughly suggested coefficients and directions.

## Data Availability

All data produced in the present study are available upon reasonable request to the authors.

## Acknowledgements

Data and/or research tools used in the preparation of this manuscript were obtained and analyzed from the controlled access datasets distributed from the Osteoarthritis Initiative (OAI), a data repository housed within the NIMH Data Archive (NDA). OAI is a collaborative informatics system created by the National Institute of Mental Health and the National Institute of Arthritis, Musculoskeletal and Skin Diseases (NIAMS) to provide a worldwide resource to quicken the pace of biomarker identification, scientific investigation and OA drug development. Dataset identifier(s): NIMH Data Archive Collection ID: 2343.

## Author Contributions

TF. L., M. N., and H. Z. designed the study. M. N. supervised the imaging analyses and H. Z. supervised the statistical analyses. TF. L. and TY. L. analyzed the data. B. C., C. H., Z. S. and Z. X. processed imaging data. TF. L., D. N., YM. G, and AE. N. provided clinical interpretations of the analyses and reviewed the paper. TF. L. and TY. L. wrote the manuscript with feedback from all authors.

## Funding Resources

Research reported in this publication was supported by the National Institutes of Health (NIH) under award number AR072013 (TY. L., B. C. and M. N.) and R01MH116527 (TF. L. and H.Z.). We also acknowledge support from NIH/NIAMS P30AR072580 (AE. N and YM. G). The content is solely the responsibility of the authors and does not necessarily represent the official views of the National Institutes of Health.

