## Supplementary file for "Charting Aging Trajectories of Knee Cartilage Thickness for Early Osteoarthritis Risk Prediction: An MRI Study from the Osteoarthritis Initiative Cohort"

September 8, 2023

**This PDF file includes**:

Supplementary Text

Tables S1 to S17

Figures S1 to S5

**Other Supplementary Information for this manuscript includes the following:**

A movie to show population-level local dynamic changes of rOA progression

### Supplementary Note

##### Alternative models of the FKTR charts

Based on 2D maps of cartilage thickness for all individuals in the common coordinate system, we have constructed the FKTR charts using the statistical model M1. To evaluate and make a comparison of model performance, alternative models M2

$$y_{i,v}\left(t_{i,j}, b_{i,j}\right) = \beta_{0,v} g_i + \beta_{1,v} b_{i,j} g_i + \beta_{2,v} g_i t_{i,j} + \beta_{3,v} R_i + \beta_{4,v} h_{i,j} + \beta_{5,v} t_{i,j} + \beta_{6,v} b_{i,j} + \beta_{7,v} b_{i,j} t_{i,j} + \alpha_{i,v} + e_{i,v}(t_{i,j})$$

and M3

$$y_{i,v}\left(t_{i,j}, b_{i,j}\right) = f_v\left(t_{i,j}\right) + \beta_{0,v} g_i + \beta_{1,v} b_{i,j} g_i + \beta_{2,v} g_i t_{i,j} + \beta_{3,v} R_i + \beta_{4,v} h_{i,j} + \beta_{6,v} b_{i,j} + \beta_{7,v} b_{i,j} t_{i,j} + \alpha_{i,v} + e_{i,v}(t_{i,j})$$

were also fitted. Model M2 only accounts for a linear relationship; on the other hand, model M3 does not incorporate the interactions between age and BMI. These models will highlight the advantages of considering nonlinear relationships and interaction effects in our models. The Akaike Information Criterion (AIC) (1) criterion and Generalized Cross-validation (GCV) errors following Ye (1998) (2) were used to compare the model performance. The AIC and GCV of models M1, M2, and M3 were shown in **Supplementary Table 1** for fitting the thickness of the knee cartilage on the left and right sides in 12 ROIs, as well as the average for the femoral and tibial regions. We found that model M1 typically provided the best fit, evidenced by its lowest GCVs and AICs. While M3 performed nearly as well as M1, both M1 and M3 significantly outperformed M2.

##### Local centile scores (Method F : controlling only the fixed effects from confounding covariates)

The proposed LCS is an adaptation of (3), where we use the GAMM model to replace the Generalized Additive Mixed Model for Location, Scale and Shape (GAMLSS) (4) model. To formulate, for a scan from the new subject *n* + 1, the LCS, represented by $LP_{n+1,v}$, can be formulated as $LP_{n+1,v} = \Pr(y_{n+1,v} - \hat{y}_{n+1,v} < y_v - \hat{y}_v)$, where $y_v$ and $y_{n+1,v}$ are cartilage thickness in the rOA-free population and the subject *n* + 1, and $\hat{y}_v$ and $\hat{y}_{n+1,v}$ are the predicted LCT at *v* based on Model M1 trained by rOA-free subjects . This centile score is equivalent to the p-value of a one-sided one-sample test of the Null Hypothesis ($H_0$): the distribution of the LCT at voxel *v* from the new subject is indistinguishable from that of the normal control population. The empirical estimate of *LP* is calculated as $\widehat{LP}_{n+1,v} = \sum_{i=1}^{n}(r_{n+1,v} \leq r_{i,v})/n$ where $r_{i,v} = y_{i,v} - \hat{y}_{i,v}$ are residuals by removing the fixed effect from $y_{i,v}$ , and $i = 1,2,\ldots,n$ are n normal subjects in the training dataset. The LCS follows a uniform distribution between [0,1] when $H_0$ holds. However, one limitation of the empirical estimate is that it cannot discriminate between extreme centile scores, for example, between $LP_{n+1,v}$ = *1e-3* and *1e-4*, when the sample size (n) is less than 1,000. To address this issue, we propose to use the wild bootstrap method (5) to resample the residuals 100,000 times, thereby enhancing its finite performance characteristics. The steps for computing the LCS are detailed in Algorithm 1. The wild bootstrap method has been shown to have strong theoretical properties and accurate finite-sample performance in several studies (6, 7).

---

**Algorithm 1.**
*Given the new subject n+1 with* age *t, sex $g$, race R, height h, and BMI $b$, and the FKTR atlas for each spatial location (pixel) v, and the total resampling times S=100,000, the following steps were performed.*
***Step1.*** *Calculated the residuals $r_{i,v}$ for reference (rOA-free) subjects (at baseline) $i = 1,\ldots,n$ and the residual $r_{n+1,v}$ for the new subject, by removing the fixed effect from observations $y_{n+1,v}$ and $y_{i,v}$ using the GAMM model, respectively.*
***Step2.*** *Generated $D_1,\cdots,D_S$ from the standard normal distribution. A bootstrap sample $\tilde{R}_i$ , $i = 1,\ldots,S$, were generated with $\tilde{R}_{i,v} = D_i * \tilde{r}_{i,v}$, where $\tilde{r}_{i,v}$ were random resamples (with replacement) from $\{r_{1,v}, r_{2,v}, r_{3,v}, \ldots, r_{n,v}\}$.*
***Step3.*** *Estimated the local centile score of the subject n+1 as $Lp_{n+1,v} = \sum_{i=1}^{S}(r_{n+1,v} \leq \tilde{R}_{i,v})/S$.*

---

**Local centile scores (Method R: controlling both the fixed and subject-level random effects)**

The above described methodology for LCS calculation evaluates each scan in isolation, disregarding the valuable information from historical scans of the same subjects. It is expected that incorporating the historical information can further improve the detection of the transition from normal to abnormal thicknesses and the monitoring of OA progression. To accomplish this, we generalize the baseline scan-level LCS for LCTs to the longitudinal scan level. Specifically, first we predict a the *N*+1-th scan from subject *n*+1 at age $t_{N+1}$, given its sex $g_{n+1}$, race $r_{n+1}$, height $h_{n+1,N+1}$ and BMI $b_{n+1,N+1}$ at $t_{N+1}$ and $h_{n+1,s}$, $b_{n+1,s}$, and $y_{n+1,v}(t_s, b_{n+1,s})$ at age $t_s$, $s = 1, \dots, N$. And then the LCS to assess the abnormality/extremity *N*+1-th scan from subject *n*+1 at each *v* by

$$LP_{n+1,v,N+1} = \Pr(y_{n+1,v}(t_{N+1}, b_{n+1,N+1}) - \hat{y}_{n+1,v}(t_{N+1}, b_{n+1,N+1}) < y_v(t_{N+1}, b_{n+1,N+1}) - \hat{y}_v(t_{N+1}, b_{n+1,N+1})),$$

where $y_v(t_{N+1}, b_{n+1,N+1})$ is the cartilage thickness in the rOA-free population at age $t_{N+1}$, and $\hat{y}_v(t_{N+1}, b_{n+1,N+1})$ is the predicted $y_v(t_{N+1}, b_{n+1,N+1})$ at location *v* using both the fixed effect and the random effect estimated from the GAMM model, using spline-based maximum restricted likelihood to obtain estimates of the regression coefficients and associated random-effects. Indeed, such centile is equivalent to the p-value of a one-sample test of the Null Hypothesis H0: the distribution of the LCT at location *v* from the new scan is the same as that from the rOA-free populations, conditioning on the current age, sex, race, BMI and height and historical scans with corresponding BMI and height measures. A simple empirical estimate of $LP_{n+1,v,N+1}$ is the empirical estimate

$$\widehat{LP}_{n+1,v,N+1} = \sum_{i=1}^{n} \sum_{j=1}^{N_i} (r_{n+1,v,N+1} \le r_{i,v,j}) / \sum_{i=1}^{n} N_i,$$

where $r_{i,v,j} = y_{i,v}(t_{i,j}, b_{i,j}) - \hat{y}_{i,v}(t_{i,j}, b_{i,j})$ are the residuals by removing the fixed and random effects from the follow-up scan LCT $y_{i,v}(t_{i,j}, b_{i,j})$, and $N_i$ are the number of scans (visits) of subject *i*. Similar to the previous subsection, we use a wild bootstrap method to improve the finite-sample properties of the estimate.

Although the estimated fixed effects exhibit statistical consistency with the true parameters when sample size is large, for the random effect of new subject, it is necessary to update the random effects when a new scan is included. While employing the entire model refitting is computationally intensive, we adopt a rapid incremental learning algorithm. In particular,

$$\hat{y}_{n+1,v}(t_{N+1}, b_{n+1,N+1}) = f_v(t_{N+1}, b_{n+1,N+1}) + \beta_{1,v} b_{n+1,N+1} g_{n+1} + \beta_{2,v} t_{N+1} g_{n+1} + \beta_{3,v} R_{n+1} + \beta_{4,v} h_{n+1} + \Delta_{n+1,v},$$

where the random effect

$$\begin{aligned}\Delta_{n+1,v} &= E[\alpha_{n+1,v} | y_{n+1,v}(t_k, b_{n+1,k}), k = 1,2, \dots, N] \\ &= E[\alpha_{n+1,v} | \alpha_{n+1,v} + e_{n+1,v}(t_k), k = 1,2, \dots, N] \\ &= \sigma_{1,v}^2 / (\sigma_{1,v}^2 + \sigma_{2,v}^2 / N) \sum_{k=1}^{N} [\alpha_{n+1,v} + e_{n+1,v}(t_k)] / N\end{aligned}$$

which can be estimated by $\hat{\Delta}_{n+1,v} = \sigma_{1,v}^2 / (\sigma_{1,v}^2 + \sigma_{2,v}^2 / N) \sum_{k=1}^{N} \hat{r}_{n+1,v,k} / N$, where $\sigma_{1,v}^2$ and $\sigma_{2,v}^2$ are the variance of random effect and random noise at the location *v*, respectively. This calculation avoids the necessity to refit the entire model through incremental learning.

Both Method F and Method R have distinct advantages in specific scenarios. Method R is more accurate when there are more historical scans for the predicted knee. In contrast, in situations with limited historical data, Method F may be more accurate by focusing on estimating the fixed effect only.

**Centile feature extraction**

We computed the GCS of a knee based on the mean centile function of its cumulative distribution function (CDF) of cartilage thickness spatial distribution among rOA-free partipants. Specifically, it was defined as

$$GCS = \int \Pr\big(F(x) \le F_0(x)\big)\,\pi(x)dx,$$

where the $F(\cdot)$ and $F_0(\cdot)$ represents the spatial CDF of the LCS of the target knee and reference knee (rOA-free knee), respectively,

$$F(x) \triangleq \int (v \in V)[LCS(v) \le x]\, dv.$$

In these equations, $v$ represents a spatial location, and $V$ denotes one of the 14 knee regions. The function $\pi(x)$ is a Bayesian prior defined over the interval [0,1], assigning importance to specific centiles of the LCS for prediction purposes. In this work we chose the data-driven $\pi(x)$ to maximize the variance of $Logit_{10}GCS$

$$\pi(x) = \max_{\pi} Var[Logit_{10}GCS(\pi)]\,, s.t.\pi \ge 0, \int_{x\in[0,1]} \pi(x)dx = 1.$$

We used such prior to capture data's most informative aspects of the data, enhancing predictive power. This optimization can be efficiently addressed using non-negative principal component analysis (NNPCA) (8).

---

**Algorithm 2.**

*Given aligned cartilage thickness maps of all knees i=1, 2,…n, n+1,…,m, and the region of interest V where the first n knees were from reference (rOA-free) participants, we calculated the GCS based on the following steps.*

***Step1.*** *Calculated the* $LCS(\cdot)$ *for each knee using* Method F *or* Method R.

***Step2.*** *For a sequence U of x, for example, x ∈ U = {0.001, 0.005, 0.01, 0.05… 0.1, 0.2}, calculated the CDF of the i-th knee* $F_i(x), i = 1,2, \dots m$. *Denote the length of sequence U as L.*

***Step3.*** *For x ∈ U,* $j = 1,2, \dots m$, *estimated the centile function* $\Pr\left(F_j(x) \le F_0(x)\right)$ *by* $\sum_{i=1,\dots n}[F_j(x) \le F_i(x)]/n$.

***Step4.*** *Performed NNPCA on the m* × $L$ *matrix* $\left(\Pr\left(F_j(x_k) \le F_0(x_k)\right)\right)_{j\le m, k\le L}$ *to obtain the loading of the first PC component* $\pi = (\pi_1, \dots, \pi_L)^T$. *Then estimated the GCS for the j-th knee as,*

$$GCS_j = \sum_{k=1,\dots L} [\Pr\left(F_j(x_k) \le F_0(x_k)\right)\pi_k]\,, j = 1,2, \dots m.$$

---

**Supplementary Table S1. The Akaike Information Criteria (AIC) and Generalized Cross Validation (GCV) values of models M1, M2 and M3.** Those were computed for ROIs 1-12, overall average of tibial (Tib) and femoral (Fem) regions on both the left and right knees. Text in Bold and italic: methods with minimum AICs or GCVs.

| AIC | | | | | | |
|---|---|---|---|---|---|---|
| ROI | M1-L | M1-R | M2-L | M2-R | M3-L | M3-R |
| 1 | ***-3866*** | ***-6002*** | -2860 | -4354 | -3853 | -5970 |
| 2 | ***-8031*** | ***-11561*** | -6109 | -8770 | -8022 | -11558 |
| 3 | -6608 | ***-6652*** | -4943 | -4849 | ***-6718*** | -6651 |
| 4 | -1393 | ***-650*** | -98 | 552 | ***-1439*** | -650 |
| 5 | ***-5540*** | ***-2200*** | -4316 | -1893 | -5530 | -2191 |
| 6 | -6668 | ***-5524*** | -5032 | -4174 | ***-6672*** | -5516 |
| 7 | ***2630*** | ***-388*** | 3105 | 1202 | 2631 | -383 |
| 8 | -493 | -5286 | 240 | -3074 | ***-529*** | ***-5295*** |
| 9 | 939 | ***-186*** | 1860 | 1161 | ***909*** | -174 |
| 10 | -593 | 3425 | -349 | 2716 | ***-608*** | ***3401*** |
| 11 | -5365 | -1864 | -3952 | -1435 | ***-5401*** | ***-1875*** |
| 12 | *-1980* | ***-2006*** | -1176 | -1045 | ***-1990*** | -1999 |
| Tib | -3431 | ***-3726*** | -2787 | -2946 | ***-3442*** | -3724 |
| Fem | ***-7043*** | ***-6017*** | -5816 | -5002 | -7032 | -5924 |
| GCV | | | | | | |
| ROI | M1-L | M1-R | M2-L | M2-R | M3-L | M3-R |
| 1 | ***0.0264*** | ***0.0185*** | 1.23 | 1.22 | 0.0266 | 0.0186 |
| 2 | ***0.0111*** | ***0.0063*** | 1.24 | 1.23 | 0.0113 | 0.0064 |
| 3 | ***0.0148*** | ***0.0163*** | 1.24 | 1.22 | 0.0148 | 0.0164 |
| 4 | 0.0445 | ***0.0524*** | 1.24 | 1.22 | ***0.0443*** | 0.0525 |
| 5 | ***0.0188*** | ***0.0384*** | 1.24 | 1.21 | 0.0188 | 0.0385 |
| 6 | ***0.0148*** | ***0.0200*** | 1.24 | 1.22 | 0.0148 | 0.0201 |
| 7 | ***0.103*** | ***0.0555*** | 1.23 | 1.22 | 0.0103 | 0.0555 |
| 8 | 0.0538 | ***0.0214*** | 1.24 | 1.22 | ***0.0534*** | 0.0214 |
| 9 | 0.0722 | ***0.0576*** | 1.24 | 1.22 | ***0.0721*** | 0.0577 |
| 10 | ***0.0521*** | 0.1138 | 1.23 | 1.20 | 0.0521 | ***0.1134*** |
| 11 | ***0.0194*** | 0.0412 | 1.24 | 1.22 | 0.0194 | ***0.0411*** |
| 12 | 0.0394 | ***0.0402*** | 1.23 | 1.21 | ***0.0393*** | 0.0403 |
| Tib | 0.0291 | ***0.0287*** | 1.24 | 1.22 | ***0.0290*** | 0.0288 |
| Fem | ***0.0137*** | ***0.0184*** | 1.24 | 1.22 | 0.0138 | 0.0187 |

**Supplementary Table S2. Feature names, identifiers, definitions, and clinical interpretations used in prediction tasks.**

| Category | Subcategory | Identifier[1] | Definition | Clinical interpretation |
|---|---|---|---|---|
| Demographic | Age | Age | Age | --- |
| Demographic | BMI | BMI | BMI | --- |
| Demographic | Sex | Sex | Sex | --- |
| Demographic | Height | Height | Height | --- |
| Demographic | Race | Race | Race | --- |
| Raw thickness | ROI-mean thickness | ROI1,<br>...,<br>ROI12,<br>WholeFem<br>WholeTib | Average of raw cartilage thickness in 14 regions (12 regions of interest (ROIs) , and the overall femoral and tibial | The increase indicates cartilage thickening |
| Centile | GPS | F_ROI1-<br>…<br>F_ROI12-<br>F_Fem-<br>F_Tib- | $Logit_{10}[GCS]_-$ in 14 regions (12 ROIs, and the overall femoral and tibial), which were calculated using method F (fixed effect only) | The increase indicates the reduced cartilage thickening |
| Centile | GPS | F_ROI1+<br>…<br>F_ROI12+<br>F_Fem+<br>F_Tib+ | $Logit_{10}[GCS]_+$ in 14 regions (12 ROIs, and the overall femoral and tibial), which were calculated using method F (fixed effect only) | The increase indicates cartilage thinning |
| Centile | GPS | R_ROI1-<br>…<br>R_ROI12-<br>R_Fem-<br>R_Tib- | $Logit_{10}[GCS]_-$ in 14 regions (12 ROIs, and the overall femoral and tibial), which were calculated using method R (Consider random effect) | The increase indicates the reduced cartilage thickening |
| Centile | GPS | R_ROI1+<br>…<br>R_ROI12+<br>R_Fem+<br>R_Tib+ | $Logit_{10}[GCS]_+$ in 14 regions (12 ROIs, and the overall femoral and tibial), which were calculated using method F (fixed effect only) | The increase indicates cartilage thinning |
| Centile | ICDF | F_ROI1Area<br>…<br>F_ROI12 Area<br>F_FemArea<br>F_TibArea | $\int F(x)\,\pi(x)dx$ in 14 regions (12 ROIs, and the overall femoral and tibial), where F is the CDF of spatial distribution of the LCS, and LCS were calculated using method F | The increase indicates cartilage thinning |
| Centile | ICDF | R_ROI1Area<br>…<br>R_ROI12Area<br>R_FemArea<br>R_TibArea | Same as the above " F_*Area" except that LCS were calculated using method R | The increase indicates cartilage thinning |

[1] The identifiers of features were used in the SHAP summary plots, boxplots, or nomograms.

**Supplementary Table S3. A comparison of 22 predictive methods on the test data, using all demographic, raw cartilage thickness and centile features, in classifying KLG = 2 and KLG < 2.** Models were evaluated through the accuracy (ACC), F1 score, AUC, recall, precision and their standard deviations. "RF" indicates performing feature selection based on their random forest permutation importance; "GLMBoost" indicates performing feature selection based on GLMBoost importance. The optimal F1 and AUC were highlighted in bold texts.

| Feature Selection | Baseline Model | ACC | Precision (KLG = 2) | Precision (KLG < 2) | Recall (KLG = 2) | Recall (KLG < 2) | F1 | AUC |
|---|---|---|---|---|---|---|---|---|
| GLMBoost | BayesGLM | 0.774 (0.002) | 0.722 (0.003) | 0.813 (0.001) | 0.748 (0.003) | 0.793 (0.004) | 0.769 (0.002) | 0.843 (0.002) |
| GLMBoost | GLMBoost | 0.769 (0.004) | 0.711 (0.005) | 0.815 (0.004) | 0.755 (0.005) | 0.779 (0.004) | 0.764 (0.004) | 0.838 (0.001) |
| GLMBoost | Glmnet | 0.779 (0.005) | 0.760 (0.007) | 0.790 (0.005) | 0.690 (0.008) | 0.843 (0.005) | 0.769 (0.005) | 0.844 (0.002) |
| GLMBoost | HDDA | 0.728 (0.006) | 0.682 (0.008) | 0.759 (0.006) | 0.658 (0.013) | 0.779 (0.009) | 0.719 (0.006) | 0.774 (0.002) |
| GLMBoost | LDA | 0.780 (0.003) | 0.769 (0.006) | 0.786 (0.002) | 0.678 (0.004) | 0.852 (0.005) | 0.769 (0.003) | 0.845 (0.002) |
| GLMBoost | LightGBM | 0.874 (0.004) | 0.876 (0.007) | 0.873 (0.002) | 0.816 (0.003) | 0.916 (0.006) | 0.870 (0.004) | 0.938 (0.003) |
| GLMBoost | Naïve Bayes | 0.745 (0.002) | 0.730 (0.005) | 0.754 (0.001) | 0.623 (0.003) | 0.834 (0.005) | 0.732 (0.002) | 0.802 (0.004) |
| GLMBoost | NNet | 0.754 (0.002) | 0.701 (0.004) | 0.794 (0.002) | 0.720 (0.005) | 0.779 (0.005) | 0.748 (0.002) | 0.826 (0.003) |
| GLMBoost | RF | 0.857 (0.003) | 0.894 (0.003) | 0.837 (0.004) | 0.746 (0.007) | 0.935 (0.002) | 0.848 (0.003) | 0.926 (0.001) |
| GLMBoost | SVM | 0.779 (0.005) | 0.762 (0.009) | 0.790 (0.003) | 0.688 (0.005) | 0.845 (0.008) | 0.770 (0.005) | 0.844 (0.002) |
| GLMBoost | XGB | 0.763 (0.004) | 0.714 (0.006) | 0.800 (0.004) | 0.725 (0.007) | 0.790 (0.007) | 0.757 (0.004) | 0.834 (0.003) |
| RF | BayesGLM | 0.763 (0.003) | 0.708 (0.003) | 0.805 (0.004) | 0.739 (0.007) | 0.781 (0.003) | 0.758 (0.003) | 0.830 (0.001) |
| RF | GLMBoost | 0.751 (0.004) | 0.689 (0.004) | 0.802 (0.004) | 0.740 (0.007) | 0.759 (0.003) | 0.747 (0.004) | 0.818 (0.001) |
| RF | Glmnet | 0.771 (0.002) | 0.758 (0.002) | 0.779 (0.003) | 0.667 (0.006) | 0.846 (0.003) | 0.760 (0.002) | 0.830 (0.002) |
| RF | HDDA | 0.661 (0.001) | 0.637 (0.006) | 0.672 (0.002) | 0.449 (0.011) | 0.815 (0.009) | 0.632 (0.002) | 0.672 (0.009) |
| RF | LDA | 0.770 (0.002) | 0.764 (0.005) | 0.773 (0.001) | 0.652 (0.002) | 0.854 (0.004) | 0.758 (0.002) | 0.830 (0.001) |
| RF | LightGBM | 0.892 (0.002) | 0.901 (0.004) | 0.885 (0.002) | 0.832 (0.003) | 0.934 (0.003) | **0.887 (0.003)** | **0.953 (0.001)** |
| RF | Naïve Bayes | 0.696 (0.004) | 0.696 (0.007) | 0.696 (0.004) | 0.487 (0.010) | 0.846 (0.005) | 0.668 (0.005) | 0.759 (0.005) |
| RF | NNet | 0.625 (0.010) | 0.551 (0.017) | 0.689 (0.019) | 0.591 (0.072) | 0.649 (0.065) | 0.617 (0.007) | 0.655 (0.012) |
| RF | RF | 0.885 (0.001) | 0.923 (0.003) | 0.865 (0.002) | 0.793 (0.004) | 0.951 (0.003) | 0.879 (0.001) | 0.943 (0.001) |
| RF | SVM | 0.769 (0.002) | 0.758 (0.003) | 0.776 (0.003) | 0.661 (0.005) | 0.848 (0.002) | 0.758 (0.003) | 0.830 (0.001) |
| RF | XGB | 0.757 (0.004) | 0.709 (0.005) | 0.792 (0.005) | 0.713 (0.009) | 0.789 (0.006) | 0.751 (0.004) | 0.824 (0.004) |

**Supplementary Table S4. A comparison of 22 predictive methods on the validation data, using all demographic, raw cartilage thickness and centile features, in classifying KLG = 2 and KLG < 2.** Models were evaluated through the accuracy (ACC), F1 score, AUC, recall, precision and their standard deviations. "RF" indicates performing feature selection based on their random forest permutation importance; "GLMBoost" means performing feature selection based on GLMBoost importance. The optimal F1 and AUC were highlighted in bold texts.

| **Feature Selection** | **Baseline Model** | **ACC** | **Precision (KLG = 2)** | **Precision (KLG < 2)** | **Recall (KLG = 2)** | **Recall (KLG < 2)** | **F1** | **AUC** |
|---|---|---|---|---|---|---|---|---|
| GLMBoost | BayesGLM | 0.778 (0.002) | 0.726 (0.002) | 0.818 (0.002) | 0.754 (0.003) | 0.794 (0.002) | 0.773 (0.002) | 0.851 (0.001) |
| GLMBoost | GLMBoost | 0.768 (0.002) | 0.711 (0.003) | 0.815 (0.002) | 0.755 (0.002) | 0.778 (0.002) | 0.764 (0.002) | 0.838 (0.001) |
| GLMBoost | Glmnet | 0.784 (0.002) | 0.766 (0.003) | 0.794 (0.002) | 0.696 (0.003) | 0.847 (0.002) | 0.774 (0.002) | 0.852 (0.001) |
| GLMBoost | HDDA | 0.730 (0.003) | 0.686 (0.004) | 0.760 (0.004) | 0.656 (0.007) | 0.783 (0.003) | 0.721 (0.003) | 0.784 (0.004) |
| GLMBoost | LDA | 0.784 (0.001) | 0.774 (0.002) | 0.790 (0.001) | 0.685 (0.002) | 0.856 (0.002) | 0.774 (0.001) | 0.852 (0.001) |
| GLMBoost | LightGBM | 0.874 (0.001) | 0.877 (0.003) | 0.872 (0.001) | 0.814 (0.002) | 0.917 (0.002) | 0.869 (0.001) | 0.938 (0.001) |
| GLMBoost | Naïve Bayes | 0.749 (0.001) | 0.736 (0.003) | 0.757 (0.002) | 0.628 (0.005) | 0.837 (0.004) | 0.736 (0.001) | 0.814 (0.001) |
| GLMBoost | NNet | 0.756 (0.002) | 0.705 (0.003) | 0.795 (0.003) | 0.720 (0.006) | 0.778 (0.008) | 0.749 (0.003) | 0.830 (0.002) |
| GLMBoost | RF | 0.854 (0.002) | 0.894 (0.003) | 0.834 (0.002) | 0.741 (0.004) | 0.935 (0.001) | 0.846 (0.003) | 0.926 (0.001) |
| GLMBoost | SVM | 0.783 (0.001) | 0.768 (0.001) | 0.792 (0.002) | 0.690 (0.003) | 0.850 (0.001) | 0.773 (0.001) | 0.851 (0.001) |
| GLMBoost | XGB | 0.767 (0.001) | 0.720 (0.003) | 0.801 (0.002) | 0.726 (0.005) | 0.796 (0.003) | 0.761 (0.002) | 0.837 (0.002) |
| RF | BayesGLM | 0.766 (0.001) | 0.711 (0.001) | 0.808 (0.002) | 0.742 (0.002) | 0.783 (0.001) | 0.761 (0.001) | 0.832 (0.001) |
| RF | GLMBoost | 0.754 (0.003) | 0.693 (0.003) | 0.803 (0.002) | 0.740 (0.003) | 0.763 (0.002) | 0.749 (0.003) | 0.821 (0.002) |
| RF | Glmnet | 0.772 (0.001) | 0.758 (0.000) | 0.780 (0.001) | 0.670 (0.002) | 0.846 (0.001) | 0.762 (0.001) | 0.832 (0.001) |
| RF | HDDA | 0.663 (0.003) | 0.639 (0.006) | 0.674 (0.002) | 0.452 (0.006) | 0.815 (0.004) | 0.633 (0.004) | 0.678 (0.001) |
| RF | LDA | 0.772 (0.001) | 0.766 (0.002) | 0.776 (0.001) | 0.658 (0.002) | 0.855 (0.001) | 0.761 (0.002) | 0.832 (0.001) |
| RF | LightGBM | 0.893 (0.001) | 0.905 (0.002) | 0.886 (0.001) | 0.834 (0.003) | 0.937 (0.001) | **0.889 (0.001)** | **0.953 (0.001)** |
| RF | Naïve Bayes | 0.700 (0.002) | 0.705 (0.002) | 0.698 (0.003) | 0.490 (0.007) | 0.852 (0.001) | 0.673 (0.003) | 0.772 (0.003) |
| RF | NNet | 0.612 (0.008) | 0.536 (0.009) | 0.677 (0.007) | 0.573 (0.013) | 0.640 (0.009) | 0.604 (0.009) | 0.641 (0.010) |
| RF | RF | 0.887 (0.000) | 0.924 (0.001) | 0.866 (0.001) | 0.796 (0.002) | 0.952 (0.001) | 0.881 (0.001) | 0.947 (0.001) |
| RF | SVM | 0.771 (0.002) | 0.761 (0.003) | 0.777 (0.001) | 0.661 (0.002) | 0.850 (0.002) | 0.760 (0.002) | 0.832 (0.001) |
| RF | XGB | 0.761 (0.001) | 0.713 (0.001) | 0.795 (0.002) | 0.718 (0.003) | 0.791 (0.002) | 0.754 (0.001) | 0.825 (0.002) |

**Supplementary Table S5. A comparison of prediction performance among different feature combinations in classifying KLG = 2 and KLG < 2.** Only the one among the 22 predictive models with the highest F1 score was shown for each feature combination. Models were evaluated through the accuracy (ACC), F1 score, AUC, recall, precision and their standard deviations. “RF” indicates performing feature selection based on their random forest permutation importance; “GLMBoost” means performing feature selection based on GLMBoost importance. The optimal F1 and AUC were highlighted in bold texts.

| Test dataset | | | | | | | | | |
|---|---|---|---|---|---|---|---|---|---|
| **Feature Combination** | **Feature Selection** | **Baseline Model** | **ACC** | **Precision (KLG = 2)** | **Precision (KLG < 2)** | **Recall (KLG = 2)** | **Recall (KLG < 2)** | **F1** | **AUC** |
| Demographic Raw Centile | RF | LightGBM | 0.892 (0.002) | 0.901 (0.004) | 0.885 (0.002) | 0.832 (0.003) | 0.934 (0.003) | **0.887 (0.003)** | **0.953 (0.001)** |
| Demographic Raw | Glmboost | LightGBM | 0.727 (0.005) | 0.670 (0.006) | 0.770 (0.005) | 0.687 (0.008) | 0.756 (0.006) | 0.721 (0.005) | 0.797 (0.002) |
| Raw | Glmboost | LightGBM | 0.700 (0.004) | 0.642 (0.005) | 0.743 (0.004) | 0.645 (0.007) | 0.740 (0.005) | 0.692 (0.004) | 0.761 (0.002) |
| Raw Centile | Glmboost | SVM | 0.770 (0.001) | 0.755 (0.002) | 0.779 (0.001) | 0.668 (0.002) | 0.844 (0.002) | 0.760 (0.001) | 0.835 (0.001) |
| **Validation dataset** | | | | | | | | | |
| **Feature Combination** | **Feature Selection** | **Baseline Model** | **ACC** | **Precision (KLG = 2)** | **Precision (KLG < 2)** | **Recall (KLG = 2)** | **Recall (KLG < 2)** | **F1** | **AUC** |
| Demographic Raw Centile | RF | LightGBM | 0.893 (0.001) | 0.905 (0.002) | 0.886 (0.001) | 0.834 (0.003) | 0.937 (0.001) | **0.889 (0.001)** | **0.953 (0.001)** |
| Demographic Raw | Glmboost | LightGBM | 0.728 (0.003) | 0.672 (0.004) | 0.770 (0.003) | 0.686 (0.004) | 0.758 (0.004) | 0.721 (0.003) | 0.798 (0.001) |
| Raw | Glmboost | RF | 0.709 (0.002) | 0.681 (0.003) | 0.725 (0.001) | 0.577 (0.002) | 0.802 (0.002) | 0.693 (0.001) | 0.761 (0.001) |
| Raw Centile | Glmboost | SVM | 0.773 (0.003) | 0.760 (0.004) | 0.781 (0.003) | 0.670 (0.005) | 0.847 (0.002) | 0.763 (0.003) | 0.841 (0.001) |

**Supplementary Table S6. A comparison of 22 predictive methods on the test data, using all demographic, raw cartilage thickness and centile features, in classifying KLG2 knees with and without progression into KLG3 within 48 months.** Models were evaluated through the accuracy (ACC), F1 score, AUC, recall, precision and their standard deviations. "RF" indicates performing feature selection based on their random forest permutation importance; "GLMBoost" indicates performing feature selection using GLMBoost importance. The optimal F1 and AUC were highlighted in bold texts.

| Feature Selection | Baseline Model | ACC | Precision (Case) | Precision (Control) | Recall (Case) | Recall (Control) | F1 | AUC |
|---|---|---|---|---|---|---|---|---|
| GLMBoost | BayesGLM | 0.695 (0.005) | 0.317 (0.007) | 0.887 (0.003) | 0.588 (0.008) | 0.718 (0.005) | 0.603 (0.006) | 0.709 (0.003) |
| GLMBoost | GLMBoost | 0.691 (0.010) | 0.321 (0.012) | 0.895 (0.005) | 0.629 (0.017) | 0.704 (0.010) | 0.607 (0.011) | 0.715 (0.004) |
| GLMBoost | Glmnet | 0.827 (0.004) | 0.577 (0.036) | 0.843 (0.003) | 0.188 (0.015) | 0.969 (0.003) | 0.592 (0.012) | 0.719 (0.002) |
| GLMBoost | HDDA | 0.760 (0.014) | 0.326 (0.037) | 0.847 (0.008) | 0.298 (0.043) | 0.863 (0.015) | 0.583 (0.022) | 0.590 (0.025) |
| GLMBoost | LDA | 0.825 (0.004) | 0.544 (0.023) | 0.852 (0.002) | 0.256 (0.012) | 0.952 (0.003) | 0.624 (0.009) | 0.716 (0.004) |
| GLMBoost | LightGBM | 0.907 (0.005) | 0.721 (0.020) | 0.954 (0.004) | 0.798 (0.020) | 0.931 (0.008) | 0.850 (0.008) | 0.957 (0.006) |
| GLMBoost | Naïve Bayes | 0.788 (0.010) | 0.382 (0.032) | 0.846 (0.003) | 0.259 (0.020) | 0.906 (0.013) | 0.591 (0.011) | 0.639 (0.011) |
| GLMBoost | NNet | 0.586 (0.031) | 0.227 (0.017) | 0.850 (0.008) | 0.526 (0.021) | 0.559 (0.092) | 0.494 (0.042) | 0.589 (0.021) |
| GLMBoost | RF | 0.921 (0.006) | 0.920 (0.021) | 0.921 (0.005) | 0.618 (0.028) | 0.987 (0.004) | 0.846 (0.013) | 0.937 (0.007) |
| GLMBoost | SVM | 0.817 (0.003) | 0.460 (0.161) | 0.820 (0.002) | 0.022 (0.012) | 0.994 (0.004) | 0.470 (0.011) | 0.689 (0.029) |
| GLMBoost | XGB | 0.772 (0.007) | 0.348 (0.014) | 0.847 (0.002) | 0.289 (0.011) | 0.879 (0.009) | 0.589 (0.006) | 0.630 (0.014) |
| RF | BayesGLM | 0.696 (0.008) | 0.317 (0.012) | 0.885 (0.006) | 0.580 (0.025) | 0.722 (0.006) | 0.603 (0.010) | 0.692 (0.007) |
| RF | GLMBoost | 0.689 (0.007) | 0.312 (0.010) | 0.885 (0.005) | 0.587 (0.019) | 0.712 (0.004) | 0.598 (0.009) | 0.695 (0.009) |
| RF | Glmnet | 0.821 (0.004) | 0.550 (0.062) | 0.830 (0.001) | 0.095 (0.004) | 0.982 (0.005) | 0.531 (0.004) | 0.697 (0.012) |
| RF | HDDA | 0.771 (0.017) | 0.345 (0.024) | 0.845 (0.006) | 0.275 (0.054) | 0.882 (0.032) | 0.583 (0.013) | 0.611 (0.022) |
| RF | LDA | 0.818 (0.006) | 0.504 (0.040) | 0.843 (0.003) | 0.201 (0.013) | 0.956 (0.006) | 0.592 (0.011) | 0.699 (0.008) |
| RF | LightGBM | 0.961 (0.004) | 0.886 (0.008) | 0.978 (0.003) | 0.902 (0.014) | 0.974 (0.002) | 0.935 (0.007) | **0.988 (0.003)** |
| RF | Naïve Bayes | 0.775 (0.004) | 0.344 (0.016) | 0.844 (0.003) | 0.263 (0.014) | 0.889 (0.004) | 0.582 (0.009) | 0.623 (0.006) |
| RF | NNet | 0.582 (0.050) | 0.224 (0.016) | 0.848 (0.009) | 0.520 (0.085) | 0.576 (0.097) | 0.496 (0.030) | 0.584 (0.022) |
| RF | RF | 0.966 (0.003) | 0.951 (0.010) | 0.970 (0.003) | 0.860 (0.012) | 0.990 (0.002) | **0.941 (0.006)** | 0.979 (0.002) |
| RF | SVM | 0.818 (0.002) | 0.517 (0.384) | 0.819 (0.001) | 0.008 (0.006) | 0.998 (0.002) | 0.457 (0.006) | 0.619 (0.012) |
| RF | XGB | 0.778 (0.004) | 0.353 (0.011) | 0.845 (0.001) | 0.265 (0.006) | 0.892 (0.006) | 0.585 (0.004) | 0.618 (0.017) |

**Supplementary Table S7. A comparison of 22 predictive methods on the validation data, using all demographic, raw cartilage thickness and centile features, in classifying KLG2 knees with and without progression into KLG3 within 48 months.** Models were evaluated through the accuracy (ACC), F1 score, AUC, recall, precision and their standard deviations. “RF” indicates performing feature selection based on their random forest permutation importance; “GLMBoost” indicates performing feature selection using GLMBoost importance. The optimal F1 and AUC were highlighted in bold texts..

| Feature Selection | Baseline Model | ACC | Precision (Case) | Precision (Control) | Recall (Case) | Recall (Control) | F1 | AUC |
|---|---|---|---|---|---|---|---|---|
| GLMBoost | BayesGLM | 0.710 (0.005) | 0.340 (0.006) | 0.898 (0.003) | 0.630 (0.011) | 0.727 (0.004) | 0.623 (0.005) | 0.736 (0.005) |
| GLMBoost | GLMBoost | 0.686 (0.005) | 0.318 (0.007) | 0.895 (0.004) | 0.631 (0.013) | 0.699 (0.004) | 0.604 (0.006) | 0.719 (0.004) |
| GLMBoost | Glmnet | 0.829 (0.001) | 0.595 (0.015) | 0.843 (0.001) | 0.185 (0.005) | 0.972 (0.002) | 0.592 (0.003) | 0.737 (0.003) |
| GLMBoost | HDDA | 0.760 (0.008) | 0.322 (0.014) | 0.849 (0.003) | 0.305 (0.024) | 0.861 (0.014) | 0.577 (0.007) | 0.609 (0.011) |
| GLMBoost | LDA | 0.826 (0.003) | 0.550 (0.015) | 0.852 (0.001) | 0.258 (0.008) | 0.953 (0.003) | 0.625 (0.006) | 0.740 (0.004) |
| GLMBoost | LightGBM | 0.912 (0.007) | 0.742 (0.020) | 0.956 (0.002) | 0.806 (0.010) | 0.936 (0.007) | 0.859 (0.009) | 0.959 (0.004) |
| GLMBoost | Naïve Bayes | 0.794 (0.003) | 0.407 (0.010) | 0.852 (0.002) | 0.290 (0.013) | 0.907 (0.005) | 0.607 (0.007) | 0.663 (0.003) |
| GLMBoost | NNet | 0.574 (0.014) | 0.235 (0.003) | 0.859 (0.005) | 0.576 (0.032) | 0.538 (0.025) | 0.493 (0.008) | 0.600 (0.007) |
| GLMBoost | RF | 0.924 (0.004) | 0.926 (0.013) | 0.924 (0.004) | 0.635 (0.019) | 0.989 (0.002) | 0.854 (0.008) | 0.937 (0.006) |
| GLMBoost | SVM | 0.818 (0.001) | 0.461 (0.066) | 0.822 (0.001) | 0.033 (0.006) | 0.993 (0.002) | 0.480 (0.005) | 0.689 (0.019) |
| GLMBoost | XGB | 0.780 (0.002) | 0.366 (0.009) | 0.849 (0.001) | 0.287 (0.006) | 0.890 (0.002) | 0.595 (0.005) | 0.639 (0.004) |
| RF | BayesGLM | 0.695 (0.004) | 0.315 (0.005) | 0.884 (0.003) | 0.576 (0.009) | 0.721 (0.003) | 0.601 (0.005) | 0.691 (0.006) |
| RF | GLMBoost | 0.689 (0.002) | 0.314 (0.003) | 0.888 (0.002) | 0.598 (0.006) | 0.709 (0.002) | 0.600 (0.003) | 0.698 (0.007) |
| RF | Glmnet | 0.821 (0.002) | 0.549 (0.028) | 0.831 (0.002) | 0.102 (0.012) | 0.981 (0.002) | 0.535 (0.010) | 0.699 (0.004) |
| RF | HDDA | 0.767 (0.009) | 0.336 (0.019) | 0.843 (0.004) | 0.259 (0.029) | 0.880 (0.015) | 0.571 (0.009) | 0.618 (0.009) |
| RF | LDA | 0.820 (0.003) | 0.514 (0.021) | 0.845 (0.002) | 0.211 (0.011) | 0.955 (0.003) | 0.597 (0.007) | 0.697 (0.004) |
| RF | LightGBM | 0.963 (0.003) | 0.893 (0.009) | 0.979 (0.001) | 0.906 (0.007) | 0.976 (0.002) | 0.938 (0.004) | **0.989 (0.002)** |
| RF | Naïve Bayes | 0.777 (0.002) | 0.354 (0.007) | 0.846 (0.001) | 0.274 (0.004) | 0.889 (0.003) | 0.587 (0.003) | 0.637 (0.002) |
| RF | NNet | 0.538 (0.027) | 0.215 (0.009) | 0.848 (0.004) | 0.571 (0.029) | 0.517 (0.043) | 0.474 (0.021) | 0.573 (0.013) |
| RF | RF | 0.968 (0.001) | 0.950 (0.004) | 0.971 (0.001) | 0.868 (0.003) | 0.990 (0.001) | **0.944 (0.002)** | 0.984 (0.002) |
| RF | SVM | 0.818 (0.001) | 0.374 (0.067) | 0.820 (0.000) | 0.014 (0.004) | 0.997 (0.001) | 0.463 (0.004) | 0.623 (0.015) |
| RF | XGB | 0.782 (0.004) | 0.365 (0.014) | 0.846 (0.002) | 0.267 (0.011) | 0.896 (0.003) | 0.589 (0.007) | 0.634 (0.013) |

**Supplementary Table S8. A comparison of prediction performance among different feature combinations in classifying KLG2 knees with and without progression into the KLG3 phase.** Only the one among the 22 predictive models with the highest F1 score was shown for each feature combination. Models were evaluated through the accuracy (ACC), F1 score, AUC, recall, precision and their standard deviations. "RF" indicates performing feature selection based on their random forest permutation importance; "GLMBoost" means performing feature selection based on GLMBoost importance. The optimal F1 and AUC were highlighted in bold texts.

| Test dataset | | | | | | | | | |
|---|---|---|---|---|---|---|---|---|---|
| **Feature Combination** | **Feature Selection** | **Baseline Model** | **ACC** | **Precision (KLG = 2)** | **Precision (KLG < 2)** | **Recall (KLG = 2)** | **Recall (KLG < 2)** | **F1** | **AUC** |
| Demographic Raw Centile | RF | RF | 0.966 (0.003) | 0.951 (0.010) | 0.970 (0.003) | 0.860 (0.012) | 0.990 (0.002) | **0.941 (0.006)** | **0.979 (0.002)** |
| Demographic Raw | RF | NNet | 0.771 (0.028) | 0.371 (0.055) | 0.855 (0.006) | 0.342 (0.044) | 0.866 (0.041) | 0.606 (0.020) | 0.608 (0.006) |
| Raw | RF | NNet | 0.779 (0.029) | 0.386 (0.055) | 0.854 (0.004) | 0.325 (0.038) | 0.880 (0.042) | 0.608 (0.019) | 0.600 (0.018) |
| Raw Centile | RF | Naïve Bayes | 0.770 (0.003) | 0.372 (0.006) | 0.861 (0.001) | 0.379 (0.005) | 0.858 (0.003) | 0.617 (0.003) | 0.654 (0.008) |
| **Validation dataset** | | | | | | | | | |
| **Feature Combination** | **Feature Selection** | **Baseline Model** | **ACC** | **Precision (KLG = 2)** | **Precision (KLG < 2)** | **Recall (KLG = 2)** | **Recall (KLG < 2)** | **F1** | **AUC** |
| Demographic Raw Centile | RF | RF | 0.968 (0.001) | 0.950 (0.004) | 0.971 (0.001) | 0.868 (0.003) | 0.990 (0.001) | **0.944 (0.002)** | **0.984 (0.002)** |
| Demographic Raw | GLMBoost | NNet | 0.759 (0.006) | 0.359 (0.010) | 0.857 (0.001) | 0.364 (0.010) | 0.836 (0.003) | 0.601 (0.003) | 0.611 (0.010) |
| Raw | RF | NNet | 0.770 (0.010) | 0.373 (0.015) | 0.853 (0.001) | 0.325 (0.015) | 0.860 (0.017) | 0.597 (0.005) | 0.595 (0.006) |
| Raw Centile | RF | Naïve Bayes | 0.776 (0.001) | 0.385 (0.003) | 0.863 (0.001) | 0.387 (0.004) | 0.862 (0.001) | 0.624 (0.002) | 0.661 (0.004) |

**Supplementary Table S9. A comparison of 22 predictive methods on the test data, using all demographic, raw cartilage thickness and centile features, in classifying KLG ≥ 2 and KLG ≤ 1.** Models were evaluated through the accuracy (ACC), F1 score, AUC, recall, precision and their standard deviations. “RF” indicates performing feature selection based on their random forest permutation importance; “GLMBoost” indicates performing feature selection based on GLMBoost importance. The optimal F1 and AUC were highlighted in bold texts.

| **Feature Selection** | **Baseline Model** | **ACC** | **Precision (Case)** | **Precision (Control)** | **Recall (Case)** | **Recall (Control)** | **F1** | **AUC** |
|---|---|---|---|---|---|---|---|---|
| GLMBoost | BayesGLM | 0.810 (0.004) | 0.879 (0.003) | 0.728 (0.006) | 0.792 (0.006) | 0.837 (0.004) | 0.806 (0.004) | 0.897 (0.001) |
| GLMBoost | GLMBoost | 0.796 (0.002) | 0.905 (0.003) | 0.691 (0.002) | 0.737 (0.002) | 0.883 (0.003) | 0.794 (0.002) | 0.883 (0.001) |
| GLMBoost | Glmnet | 0.818 (0.002) | 0.848 (0.003) | 0.772 (0.001) | 0.848 (0.001) | 0.772 (0.006) | 0.810 (0.002) | 0.899 (0.001) |
| GLMBoost | HDDA | 0.741 (0.002) | 0.850 (0.003) | 0.638 (0.002) | 0.691 (0.003) | 0.817 (0.004) | 0.739 (0.002) | 0.798 (0.003) |
| GLMBoost | LDA | 0.815 (0.005) | 0.857 (0.004) | 0.757 (0.006) | 0.831 (0.005) | 0.792 (0.005) | 0.809 (0.005) | 0.898 (0.002) |
| GLMBoost | LightGBM | 0.823 (0.001) | 0.876 (0.003) | 0.754 (0.001) | 0.821 (0.002) | 0.826 (0.006) | **0.818 (0.001)** | **0.900 (0.003)** |
| GLMBoost | Naïve Bayes | 0.765 (0.002) | 0.814 (0.007) | 0.698 (0.004) | 0.790 (0.008) | 0.729 (0.014) | 0.757 (0.002) | 0.838 (0.002) |
| GLMBoost | NNet | 0.682 (0.007) | 0.768 (0.019) | 0.588 (0.013) | 0.677 (0.037) | 0.690 (0.049) | 0.676 (0.007) | 0.734 (0.020) |
| GLMBoost | RF | 0.805 (0.003) | 0.839 (0.003) | 0.755 (0.004) | 0.836 (0.003) | 0.756 (0.004) | 0.796 (0.003) | 0.888 (0.002) |
| GLMBoost | SVM | 0.810 (0.005) | 0.840 (0.005) | 0.766 (0.006) | 0.846 (0.005) | 0.757 (0.009) | 0.802 (0.005) | 0.894 (0.001) |
| GLMBoost | XGB | 0.817 (0.005) | 0.868 (0.004) | 0.750 (0.007) | 0.819 (0.006) | 0.814 (0.005) | 0.812 (0.005) | 0.895 (0.003) |
| RF | BayesGLM | 0.801 (0.003) | 0.873 (0.003) | 0.716 (0.004) | 0.781 (0.003) | 0.829 (0.005) | 0.797 (0.004) | 0.888 (0.002) |
| RF | GLMBoost | 0.779 (0.003) | 0.884 (0.001) | 0.676 (0.004) | 0.727 (0.004) | 0.857 (0.001) | 0.777 (0.003) | 0.870 (0.002) |
| RF | Glmnet | 0.804 (0.002) | 0.837 (0.002) | 0.755 (0.003) | 0.837 (0.003) | 0.755 (0.004) | 0.796 (0.002) | 0.888 (0.001) |
| RF | HDDA | 0.753 (0.004) | 0.838 (0.009) | 0.660 (0.003) | 0.729 (0.007) | 0.789 (0.016) | 0.749 (0.004) | 0.812 (0.004) |
| RF | LDA | 0.805 (0.002) | 0.844 (0.003) | 0.749 (0.002) | 0.828 (0.002) | 0.770 (0.005) | 0.798 (0.002) | 0.889 (0.002) |
| RF | LightGBM | 0.811 (0.006) | 0.865 (0.007) | 0.742 (0.007) | 0.812 (0.006) | 0.810 (0.011) | 0.806 (0.006) | 0.892 (0.002) |
| RF | Naïve Bayes | 0.754 (0.003) | 0.811 (0.014) | 0.679 (0.014) | 0.770 (0.024) | 0.729 (0.033) | 0.746 (0.003) | 0.823 (0.004) |
| RF | NNet | 0.721 (0.010) | 0.808 (0.010) | 0.627 (0.018) | 0.703 (0.034) | 0.748 (0.027) | 0.716 (0.008) | 0.768 (0.011) |
| RF | RF | 0.802 (0.005) | 0.842 (0.004) | 0.744 (0.006) | 0.824 (0.004) | 0.766 (0.004) | 0.794 (0.004) | 0.882 (0.002) |
| RF | SVM | 0.805 (0.004) | 0.834 (0.005) | 0.760 (0.003) | 0.842 (0.003) | 0.748 (0.010) | 0.796 (0.004) | 0.886 (0.002) |
| RF | XGB | 0.808 (0.000) | 0.855 (0.000) | 0.745 (0.000) | 0.819 (0.000) | 0.792 (0.000) | 0.802 (0.000) | 0.881 (0.000) |

**Supplementary Table S10. A comparison of 22 predictive methods on the validation data, using all demographic, raw cartilage thickness and centile features, in classifying KLG ≥ 2 and KLG ≤ 1.** Models were evaluated through the accuracy (ACC), F1 score, AUC, recall, precision and their standard deviations. "RF" indicates performing feature selection based on their random forest permutation importance; "GLMBoost" indicates performing feature selection based on GLMBoost importance. The optimal F1 and AUC were highlighted in bold texts.

| Feature Selection | Baseline Model | ACC | Precision (Case) | Precision (Control) | Recall (Case) | Recall (Control) | F1 | AUC |
|---|---|---|---|---|---|---|---|---|
| GLMBoost | BayesGLM | 0.814 (0.001) | 0.882 (0.002) | 0.734 (0.001) | 0.797 (0.001) | 0.840 (0.003) | 0.811 (0.001) | 0.900 (0.001) |
| GLMBoost | GLMBoost | 0.797 (0.002) | 0.906 (0.002) | 0.693 (0.002) | 0.739 (0.002) | 0.885 (0.002) | 0.795 (0.002) | 0.883 (0.001) |
| GLMBoost | Glmnet | 0.820 (0.001) | 0.850 (0.001) | 0.774 (0.001) | 0.849 (0.001) | 0.775 (0.002) | 0.812 (0.001) | 0.901 (0.001) |
| GLMBoost | HDDA | 0.741 (0.001) | 0.850 (0.001) | 0.637 (0.001) | 0.690 (0.001) | 0.817 (0.002) | 0.739 (0.001) | 0.801 (0.001) |
| GLMBoost | LDA | 0.818 (0.001) | 0.859 (0.001) | 0.760 (0.002) | 0.833 (0.002) | 0.795 (0.002) | 0.811 (0.001) | **0.902 (0.001)** |
| GLMBoost | LightGBM | 0.822 (0.002) | 0.876 (0.002) | 0.752 (0.003) | 0.819 (0.003) | 0.826 (0.003) | **0.817 (0.002)** | 0.900 (0.002) |
| GLMBoost | Naïve Bayes | 0.768 (0.001) | 0.818 (0.007) | 0.702 (0.005) | 0.791 (0.009) | 0.732 (0.016) | 0.759 (0.002) | 0.843 (0.001) |
| GLMBoost | NNet | 0.677 (0.004) | 0.750 (0.006) | 0.590 (0.005) | 0.699 (0.011) | 0.640 (0.011) | 0.666 (0.004) | 0.723 (0.005) |
| GLMBoost | RF | 0.807 (0.001) | 0.841 (0.001) | 0.757 (0.001) | 0.837 (0.001) | 0.759 (0.003) | 0.799 (0.001) | 0.889 (0.001) |
| GLMBoost | SVM | 0.815 (0.002) | 0.844 (0.002) | 0.771 (0.002) | 0.849 (0.001) | 0.764 (0.003) | 0.807 (0.002) | 0.898 (0.001) |
| GLMBoost | XGB | 0.815 (0.001) | 0.868 (0.000) | 0.747 (0.002) | 0.817 (0.002) | 0.813 (0.001) | 0.810 (0.001) | 0.894 (0.001) |
| RF | BayesGLM | 0.799 (0.001) | 0.872 (0.002) | 0.714 (0.002) | 0.780 (0.002) | 0.828 (0.003) | 0.795 (0.002) | 0.888 (0.001) |
| RF | GLMBoost | 0.780 (0.001) | 0.885 (0.001) | 0.677 (0.001) | 0.727 (0.001) | 0.859 (0.001) | 0.778 (0.001) | 0.870 (0.001) |
| RF | Glmnet | 0.805 (0.001) | 0.837 (0.001) | 0.756 (0.001) | 0.838 (0.001) | 0.755 (0.001) | 0.796 (0.001) | 0.888 (0.000) |
| RF | HDDA | 0.754 (0.001) | 0.841 (0.002) | 0.660 (0.001) | 0.729 (0.002) | 0.792 (0.003) | 0.750 (0.001) | 0.820 (0.002) |
| RF | LDA | 0.806 (0.002) | 0.845 (0.002) | 0.750 (0.002) | 0.829 (0.002) | 0.771 (0.003) | 0.799 (0.002) | 0.888 (0.000) |
| RF | LightGBM | 0.811 (0.003) | 0.866 (0.003) | 0.740 (0.003) | 0.810 (0.002) | 0.812 (0.005) | 0.806 (0.003) | 0.892 (0.002) |
| RF | Naïve Bayes | 0.757 (0.004) | 0.815 (0.014) | 0.688 (0.016) | 0.776 (0.026) | 0.730 (0.034) | 0.749 (0.003) | 0.832 (0.003) |
| RF | NNet | 0.716 (0.005) | 0.804 (0.009) | 0.624 (0.007) | 0.701 (0.014) | 0.736 (0.019) | 0.710 (0.006) | 0.759 (0.007) |
| RF | RF | 0.803 (0.002) | 0.843 (0.001) | 0.746 (0.003) | 0.825 (0.003) | 0.766 (0.002) | 0.795 (0.002) | 0.884 (0.001) |
| RF | SVM | 0.804 (0.001) | 0.833 (0.002) | 0.760 (0.002) | 0.843 (0.002) | 0.745 (0.004) | 0.795 (0.002) | 0.885 (0.001) |
| RF | XGB | 0.806 (0.000) | 0.857 (0.000) | 0.740 (0.000) | 0.814 (0.000) | 0.796 (0.000) | 0.801 (0.000) | 0.885 (0.000) |

**Supplementary Table S11. A comparison of prediction performance among different feature combinations in classifying KLG ≥ 2 and KLG ≤ 1.** Only the one among the 22 predictive models with the highest F1 score was shown for each feature combination. Models were evaluated through the accuracy (ACC), F1 score, AUC, recall, precision and their standard deviations. "RF" indicates performing feature selection based on their random forest permutation importance; "GLMBoost" means performing feature selection based on GLMBoost importance. The optimal F1 and AUC were highlighted in bold texts.

| **Test dataset** | | | | | | | | | |
|---|---|---|---|---|---|---|---|---|---|
| **Feature Combination** | **Feature Selection** | **Baseline Model** | **ACC** | **Precision (KLG = 2)** | **Precision (KLG < 2)** | **Recall (KLG = 2)** | **Recall (KLG < 2)** | **F1** | **AUC** |
| Demographic Raw Centile | GLMBoost | LightGBM | 0.823 (0.001) | 0.876 (0.003) | 0.754 (0.001) | 0.821 (0.002) | 0.826 (0.006) | **0.818 (0.001)** | **0.900 (0.003)** |
| Demographic Raw | RF | LightGBM | 0.790 (0.004) | 0.867 (0.005) | 0.703 (0.005) | 0.768 (0.005) | 0.822 (0.007) | 0.786 (0.004) | 0.873 (0.001) |
| Raw | GLMBoost | HDDA | 0.783 (0.004) | 0.849 (0.004) | 0.702 (0.004) | 0.776 (0.003) | 0.793 (0.007) | 0.778 (0.004) | 0.862 (0.002) |
| Raw Centile | GLMBoost | LightGBM | 0.806 (0.002) | 0.864 (0.005) | 0.732 (0.005) | 0.803 (0.007) | 0.810 (0.009) | 0.800 (0.002) | 0.885 (0.002) |
| **Validation dataset** | | | | | | | | | |
| **Feature Combination** | **Feature Selection** | **Baseline Model** | **ACC** | **Precision (KLG = 2)** | **Precision (KLG < 2)** | **Recall (KLG = 2)** | **Recall (KLG < 2)** | **F1** | **AUC** |
| Demographic Raw Centile | GLMBoost | LightGBM | 0.822 (0.002) | 0.876 (0.002) | 0.752 (0.003) | 0.819 (0.003) | 0.826 (0.003) | **0.817 (0.002)** | **0.900 (0.002)** |
| Demographic Raw | RF | LightGBM | 0.787 (0.002) | 0.863 (0.003) | 0.700 (0.003) | 0.766 (0.003) | 0.817 (0.005) | 0.783 (0.003) | 0.871 (0.001) |
| Raw | GLMBoost | HDDA | 0.779 (0.003) | 0.847 (0.003) | 0.696 (0.003) | 0.771 (0.002) | 0.790 (0.005) | 0.774 (0.003) | 0.860 (0.001) |
| Raw Centile | RF | LightGBM | 0.804 (0.002) | 0.862 (0.003) | 0.732 (0.002) | 0.803 (0.002) | 0.806 (0.005) | 0.799 (0.002) | 0.886 (0.002) |

**Supplementary Table S12. A comparison of 22 predictive methods on the test data, using all demographic, raw cartilage thickness and centile features, in predicting rOA incidence within 48 months.** Models were evaluated through the accuracy (ACC), F1 score, AUC, recall, precision and their standard deviations. "RF" indicates performing feature selection based on their random forest permutation importance; "GLMBoost" indicates performing feature selection using GLMBoost importance. The optimal F1 and AUC were highlighted in bold texts.

| Feature Selection | Baseline Model | ACC | Precision (Case) | Precision (Control) | Recall (Case) | Recall (Control) | F1 | AUC |
|---|---|---|---|---|---|---|---|---|
| GLMBoost | BayesGLM | 0.867 (0.003) | 0.620 (0.023) | 0.888 (0.001) | 0.317 (0.007) | 0.965 (0.003) | 0.672 (0.005) | 0.810 (0.004) |
| GLMBoost | GLMBoost | 0.868 (0.002) | 0.731 (0.018) | 0.874 (0.002) | 0.201 (0.013) | 0.987 (0.001) | 0.621 (0.009) | 0.817 (0.002) |
| GLMBoost | Glmnet | 0.871 (0.001) | 0.669 (0.014) | 0.885 (0.001) | 0.291 (0.010) | 0.974 (0.002) | 0.666 (0.005) | 0.816 (0.003) |
| GLMBoost | HDDA | 0.787 (0.009) | 0.262 (0.026) | 0.865 (0.004) | 0.224 (0.033) | 0.888 (0.012) | 0.559 (0.015) | 0.560 (0.017) |
| GLMBoost | LDA | 0.866 (0.002) | 0.593 (0.013) | 0.893 (0.001) | 0.358 (0.008) | 0.956 (0.003) | **0.685 (0.004)** | 0.813 (0.002) |
| GLMBoost | LightGBM | 0.870 (0.003) | 0.680 (0.030) | 0.882 (0.002) | 0.270 (0.016) | 0.977 (0.004) | 0.657 (0.009) | 0.814 (0.003) |
| GLMBoost | Naïve Bayes | 0.853 (0.002) | 0.614 (0.050) | 0.859 (0.001) | 0.086 (0.007) | 0.990 (0.002) | 0.535 (0.006) | 0.714 (0.007) |
| GLMBoost | NNet | 0.848 (0.001) | 0.094 (0.210) | 0.849 (0.002) | 0.007 (0.015) | 0.998 (0.003) | 0.465 (0.014) | 0.530 (0.018) |
| GLMBoost | RF | 0.867 (0.003) | 0.657 (0.030) | 0.880 (0.003) | 0.252 (0.024) | 0.976 (0.003) | 0.645 (0.014) | 0.806 (0.004) |
| GLMBoost | SVM | 0.860 (0.004) | 0.650 (0.036) | 0.869 (0.003) | 0.167 (0.026) | 0.984 (0.001) | 0.594 (0.019) | 0.806 (0.003) |
| GLMBoost | XGB | 0.865 (0.003) | 0.615 (0.018) | 0.885 (0.003) | 0.295 (0.022) | 0.967 (0.002) | 0.661 (0.013) | 0.806 (0.008) |
| RF | BayesGLM | 0.867 (0.002) | 0.626 (0.014) | 0.886 (0.001) | 0.299 (0.007) | 0.968 (0.001) | 0.665 (0.005) | **0.817 (0.003)** |
| RF | GLMBoost | 0.868 (0.002) | 0.722 (0.019) | 0.874 (0.002) | 0.203 (0.011) | 0.986 (0.001) | 0.622 (0.008) | 0.815 (0.002) |
| RF | Glmnet | 0.868 (0.005) | 0.657 (0.036) | 0.882 (0.004) | 0.272 (0.027) | 0.975 (0.002) | 0.655 (0.017) | 0.817 (0.003) |
| RF | HDDA | 0.810 (0.009) | 0.308 (0.014) | 0.866 (0.004) | 0.206 (0.042) | 0.917 (0.018) | 0.568 (0.013) | 0.571 (0.015) |
| RF | LDA | 0.867 (0.004) | 0.608 (0.023) | 0.892 (0.003) | 0.346 (0.016) | 0.960 (0.002) | 0.683 (0.010) | 0.815 (0.003) |
| RF | LightGBM | 0.870 (0.003) | 0.698 (0.035) | 0.880 (0.002) | 0.249 (0.015) | 0.981 (0.004) | 0.647 (0.008) | 0.812 (0.003) |
| RF | Naïve Bayes | 0.851 (0.004) | 0.534 (0.049) | 0.867 (0.001) | 0.160 (0.009) | 0.975 (0.005) | 0.581 (0.005) | 0.737 (0.011) |
| RF | NNet | 0.848 (0.001) | 0.050 (0.112) | 0.849 (0.000) | 0.001 (0.002) | 0.999 (0.001) | 0.460 (0.002) | 0.527 (0.018) |
| RF | RF | 0.869 (0.002) | 0.651 (0.015) | 0.884 (0.004) | 0.285 (0.027) | 0.972 (0.004) | 0.661 (0.013) | 0.799 (0.004) |
| RF | SVM | 0.858 (0.001) | 0.656 (0.012) | 0.865 (0.001) | 0.135 (0.007) | 0.987 (0.001) | 0.573 (0.005) | 0.794 (0.010) |
| RF | XGB | 0.862 (0.002) | 0.601 (0.013) | 0.881 (0.002) | 0.267 (0.013) | 0.968 (0.002) | 0.646 (0.007) | 0.804 (0.011) |

**Supplementary Table S13. A comparison of 22 predictive methods on the validation data, using all demographic, raw cartilage thickness and centile features, in predicting rOA incidence within 48 months.** Models were evaluated through the accuracy (ACC), F1 score, AUC, recall, precision and their standard deviations. "RF" indicates performing feature selection based on their random forest permutation importance; "GLMBoost" indicates performing feature selection using GLMBoost importance. The optimal F1 and AUC were highlighted in bold texts..

| Feature Selection | Baseline Model | ACC | Precision (Case) | Precision (Control) | Recall (Case) | Recall (Control) | F1 | AUC |
|---|---|---|---|---|---|---|---|---|
| GLMBoost | BayesGLM | 0.871 (0.001) | 0.647 (0.007) | 0.890 (0.001) | 0.328 (0.007) | 0.968 (0.001) | 0.681 (0.004) | 0.825 (0.002) |
| GLMBoost | GLMBoost | 0.868 (0.001) | 0.730 (0.013) | 0.875 (0.001) | 0.208 (0.007) | 0.986 (0.001) | 0.625 (0.005) | 0.818 (0.002) |
| GLMBoost | Glmnet | 0.874 (0.002) | 0.693 (0.013) | 0.887 (0.001) | 0.304 (0.006) | 0.976 (0.001) | 0.676 (0.004) | **0.827 (0.001)** |
| GLMBoost | HDDA | 0.794 (0.005) | 0.291 (0.052) | 0.864 (0.001) | 0.200 (0.004) | 0.900 (0.006) | 0.544 (0.002) | 0.551 (0.001) |
| GLMBoost | LDA | 0.869 (0.001) | 0.613 (0.006) | 0.896 (0.001) | 0.374 (0.004) | 0.958 (0.001) | **0.695 (0.003)** | 0.825 (0.002) |
| GLMBoost | LightGBM | 0.871 (0.002) | 0.700 (0.020) | 0.881 (0.001) | 0.254 (0.010) | 0.980 (0.002) | 0.650 (0.006) | 0.820 (0.001) |
| GLMBoost | Naïve Bayes | 0.854 (0.001) | 0.602 (0.010) | 0.860 (0.001) | 0.095 (0.005) | 0.989 (0.000) | 0.541 (0.004) | 0.742 (0.004) |
| GLMBoost | NNet | 0.849 (0.001) | 0.257 (0.075) | 0.851 (0.001) | 0.018 (0.006) | 0.997 (0.001) | 0.475 (0.005) | 0.535 (0.012) |
| GLMBoost | RF | 0.868 (0.001) | 0.671 (0.006) | 0.880 (0.002) | 0.255 (0.018) | 0.976 (0.002) | 0.646 (0.010) | 0.811 (0.003) |
| GLMBoost | SVM | 0.862 (0.002) | 0.670 (0.013) | 0.870 (0.003) | 0.171 (0.019) | 0.986 (0.001) | 0.596 (0.013) | 0.811 (0.006) |
| GLMBoost | XGB | 0.866 (0.002) | 0.621 (0.012) | 0.885 (0.001) | 0.297 (0.008) | 0.968 (0.001) | 0.663 (0.005) | 0.813 (0.004) |
| RF | BayesGLM | 0.868 (0.002) | 0.632 (0.012) | 0.886 (0.001) | 0.305 (0.008) | 0.968 (0.001) | 0.668 (0.005) | 0.819 (0.003) |
| RF | GLMBoost | 0.868 (0.001) | 0.722 (0.008) | 0.875 (0.001) | 0.208 (0.005) | 0.986 (0.000) | 0.625 (0.003) | 0.816 (0.002) |
| RF | Glmnet | 0.869 (0.001) | 0.669 (0.009) | 0.881 (0.001) | 0.264 (0.006) | 0.977 (0.001) | 0.652 (0.004) | 0.818 (0.002) |
| RF | HDDA | 0.806 (0.008) | 0.338 (0.052) | 0.867 (0.003) | 0.216 (0.034) | 0.911 (0.015) | 0.559 (0.015) | 0.569 (0.012) |
| RF | LDA | 0.866 (0.001) | 0.600 (0.008) | 0.892 (0.001) | 0.350 (0.005) | 0.958 (0.001) | 0.683 (0.003) | 0.819 (0.003) |
| RF | LightGBM | 0.867 (0.001) | 0.682 (0.014) | 0.877 (0.001) | 0.233 (0.009) | 0.980 (0.002) | 0.636 (0.005) | 0.817 (0.002) |
| RF | Naïve Bayes | 0.853 (0.002) | 0.548 (0.019) | 0.867 (0.001) | 0.161 (0.008) | 0.976 (0.002) | 0.583 (0.005) | 0.758 (0.007) |
| RF | NNet | 0.848 (0.001) | 0.023 (0.019) | 0.849 (0.000) | 0.000 (0.000) | 0.999 (0.001) | 0.459 (0.000) | 0.515 (0.006) |
| RF | RF | 0.868 (0.002) | 0.664 (0.025) | 0.881 (0.001) | 0.260 (0.008) | 0.976 (0.003) | 0.649 (0.005) | 0.810 (0.003) |
| RF | SVM | 0.860 (0.001) | 0.666 (0.021) | 0.866 (0.002) | 0.141 (0.012) | 0.988 (0.001) | 0.576 (0.009) | 0.802 (0.005) |
| RF | XGB | 0.864 (0.002) | 0.620 (0.013) | 0.880 (0.001) | 0.260 (0.007) | 0.971 (0.001) | 0.645 (0.005) | 0.809 (0.001) |

**Supplementary Table S14. A comparison of prediction performance among different feature combinations in predicting rOA incidence within 48 months.** Only the one among the 22 predictive models with the highest F1 score was shown for each feature combination. Models were evaluated through the accuracy (ACC), F1 score, AUC, recall, precision and their standard deviations. "RF" indicates performing feature selection based on their random forest permutation importance; "GLMBoost" means performing feature selection based on GLMBoost importance. The optimal F1 and AUC were highlighted in bold texts.

| Test dataset | | | | | | | | | |
|---|---|---|---|---|---|---|---|---|---|
| **Feature Combination** | **Feature Selection** | **Baseline Model** | **ACC** | **Precision (KLG = 2)** | **Precision (KLG < 2)** | **Recall (KLG = 2)** | **Recall (KLG < 2)** | **F1** | **AUC** |
| KLG Demographic Raw Centile | GLMBoost | LDA | 0.866 (0.002) | 0.593 (0.013) | 0.893 (0.001) | 0.358 (0.008) | 0.956 (0.003) | **0.685 (0.004)** | **0.813 (0.002)** |
| KLG Demographic Raw | RF | HDDA | 0.834 (0.004) | 0.404 (0.027) | 0.869 (0.003) | 0.201 (0.017) | 0.947 (0.003) | 0.587 (0.011) | 0.771 (0.003) |
| KLG Demographic | GLMBoost | LDA | 0.853 (0.003) | 0.550 (0.032) | 0.866 (0.001) | 0.153 (0.009) | 0.978 (0.002) | 0.579 (0.008) | 0.775 (0.002) |
| KLG Raw | RF | HDDA | 0.835 (0.002) | 0.402 (0.009) | 0.868 (0.002) | 0.191 (0.014) | 0.949 (0.003) | 0.583 (0.007) | 0.732 (0.007) |
| KLG Raw Centile | GLMBoost | BayesGLM | 0.729 (0.008) | 0.595 (0.024) | 0.763 (0.003) | 0.386 (0.009) | 0.883 (0.011) | 0.643 (0.008) | 0.709 (0.007) |
| **Validation dataset** | | | | | | | | | |
| **Feature Combination** | **Feature Selection** | **Baseline Model** | **ACC** | **Precision (KLG = 2)** | **Precision (KLG < 2)** | **Recall (KLG = 2)** | **Recall (KLG < 2)** | **F1** | **AUC** |
| KLG Demographic Raw Centile | GLMBoost | LDA | 0.869 (0.001) | 0.613 (0.006) | 0.896 (0.001) | 0.374 (0.004) | 0.958 (0.001) | **0.695 (0.003)** | **0.825 (0.002)** |
| KLG Demographic Raw | RF | HDDA | 0.836 (0.002) | 0.418 (0.010) | 0.871 (0.002) | 0.212 (0.013) | 0.947 (0.004) | 0.594 (0.006) | 0.776 (0.002) |
| KLG Demographic | GLMBoost | LDA | 0.851 (0.001) | 0.534 (0.015) | 0.866 (0.001) | 0.150 (0.006) | 0.977 (0.001) | 0.576 (0.005) | 0.774 (0.002) |
| KLG Raw | RF | HDDA | 0.834 (0.005) | 0.413 (0.029) | 0.869 (0.002) | 0.195 (0.013) | 0.948 (0.006) | 0.583 (0.009) | 0.742 (0.006) |
| KLG Raw Centile | GLMBoost | BayesGLM | 0.747 (0.003) | 0.637 (0.004) | 0.775 (0.003) | 0.416 (0.009) | 0.894 (0.001) | 0.667 (0.005) | 0.746 (0.003) |

**Supplementary Table S15. A comparison of 22 predictive methods on the test data, using only the raw cartilage thickness and centile features, in predicting rOA initial transition within 48 months.** Models were evaluated through the accuracy (ACC), F1 score, AUC, recall, precision and their standard deviations. “RF” indicates performing feature selection based on their random forest permutation importance; “GLMBoost” indicates performing feature selection using GLMBoost importance. The optimal F1 and AUC were highlighted in bold texts.

| Feature Selection | Baseline Model | ACC | Precision (Case) | Precision (Control) | Recall (Case) | Recall (Control) | F1 | AUC |
|---|---|---|---|---|---|---|---|---|
| GLMBoost | BayesGLM | 0.539 (0.033) | 0.538 (0.033) | 0.540 (0.033) | 0.561 (0.036) | 0.517 (0.050) | 0.539 (0.033) | 0.529 (0.019) |
| GLMBoost | GLMBoost | 0.600 (0.035) | 0.605 (0.040) | 0.596 (0.031) | 0.583 (0.042) | 0.617 (0.059) | 0.599 (0.035) | 0.574 (0.031) |
| GLMBoost | Glmnet | 0.626 (0.040) | 0.641 (0.046) | 0.614 (0.035) | 0.574 (0.042) | 0.678 (0.042) | 0.625 (0.040) | 0.579 (0.050) |
| GLMBoost | HDDA | 0.528 (0.059) | 0.523 (0.058) | 0.534 (0.061) | 0.539 (0.118) | 0.517 (0.039) | 0.527 (0.059) | 0.565 (0.036) |
| GLMBoost | LDA | 0.546 (0.018) | 0.543 (0.017) | 0.549 (0.019) | 0.578 (0.033) | 0.513 (0.039) | 0.545 (0.018) | 0.519 (0.023) |
| GLMBoost | LightGBM | 0.583 (0.043) | 0.585 (0.044) | 0.580 (0.043) | 0.561 (0.068) | 0.604 (0.042) | 0.582 (0.044) | 0.569 (0.027) |
| GLMBoost | Naïve Bayes | 0.596 (0.014) | 0.591 (0.017) | 0.602 (0.012) | 0.626 (0.032) | 0.565 (0.049) | 0.595 (0.016) | 0.585 (0.013) |
| GLMBoost | NNet | 0.502 (0.062) | 0.510 (0.080) | 0.498 (0.052) | 0.422 (0.063) | 0.583 (0.130) | 0.497 (0.059) | 0.542 (0.042) |
| GLMBoost | RF | 0.598 (0.038) | 0.606 (0.042) | 0.591 (0.036) | 0.561 (0.042) | 0.626 (0.042) | 0.595 (0.037) | 0.594 (0.034) |
| GLMBoost | SVM | 0.559 (0.024) | 0.558 (0.026) | 0.560 (0.023) | 0.574 (0.039) | 0.543 (0.061) | 0.558 (0.024) | 0.537 (0.023) |
| GLMBoost | XGB | 0.533 (0.066) | 0.533 (0.064) | 0.532 (0.068) | 0.526 (0.068) | 0.539 (0.074) | 0.532 (0.066) | 0.546 (0.034) |
| RF | BayesGLM | 0.557 (0.012) | 0.554 (0.008) | 0.560 (0.018) | 0.578 (0.045) | 0.535 (0.025) | 0.556 (0.011) | 0.556 (0.032) |
| RF | GLMBoost | 0.615 (0.029) | 0.622 (0.034) | 0.610 (0.027) | 0.591 (0.039) | 0.639 (0.050) | 0.615 (0.029) | 0.617 (0.034) |
| RF | Glmnet | 0.659 (0.027) | 0.679 (0.028) | 0.643 (0.028) | 0.600 (0.048) | 0.700 (0.047) | **0.653 (0.036)** | **0.639 (0.021)** |
| RF | HDDA | 0.565 (0.032) | 0.556 (0.030) | 0.581 (0.039) | 0.652 (0.072) | 0.478 (0.074) | 0.560 (0.032) | 0.566 (0.043) |
| RF | LDA | 0.574 (0.039) | 0.569 (0.032) | 0.581 (0.047) | 0.604 (0.068) | 0.543 (0.027) | 0.573 (0.038) | 0.566 (0.026) |
| RF | LightGBM | 0.528 (0.038) | 0.532 (0.043) | 0.526 (0.035) | 0.491 (0.050) | 0.565 (0.063) | 0.527 (0.038) | 0.549 (0.051) |
| RF | Naïve Bayes | 0.602 (0.023) | 0.589 (0.027) | 0.623 (0.023) | 0.687 (0.045) | 0.517 (0.071) | 0.598 (0.025) | 0.589 (0.039) |
| RF | NNet | 0.530 (0.036) | 0.529 (0.038) | 0.535 (0.041) | 0.578 (0.094) | 0.483 (0.100) | 0.526 (0.035) | 0.540 (0.032) |
| RF | RF | 0.554 (0.040) | 0.556 (0.043) | 0.553 (0.038) | 0.552 (0.019) | 0.557 (0.068) | 0.554 (0.040) | 0.610 (0.028) |
| RF | SVM | 0.567 (0.024) | 0.568 (0.018) | 0.568 (0.030) | 0.557 (0.068) | 0.578 (0.025) | 0.567 (0.024) | 0.548 (0.023) |
| RF | XGB | 0.561 (0.064) | 0.562 (0.070) | 0.560 (0.059) | 0.548 (0.083) | 0.574 (0.070) | 0.560 (0.064) | 0.583 (0.063) |

**Supplementary Table S16. A comparison of 22 predictive methods on the validation data, using all demographic, raw cartilage thickness and centile features, in predicting rOA initial transition within 48 months.** Models were evaluated through the accuracy (ACC), F1 score, AUC, recall, precision and their standard deviations. "RF" indicates performing feature selection based on their random forest permutation importance; "GLMBoost" indicates performing feature selection using GLMBoost importance. The optimal F1 and AUC were highlighted in bold texts..

| Feature Selection | Baseline Model | ACC | Precision (Case) | Precision (Control) | Recall (Case) | Recall (Control) | F1 | AUC |
|---|---|---|---|---|---|---|---|---|
| GLMBoost | BayesGLM | 0.653 (0.016) | 0.648 (0.014) | 0.659 (0.019) | 0.671 (0.023) | 0.635 (0.014) | 0.653 (0.016) | **0.707 (0.019)** |
| GLMBoost | GLMBoost | 0.649 (0.012) | 0.657 (0.015) | 0.644 (0.010) | 0.628 (0.006) | 0.670 (0.018) | 0.649 (0.012) | 0.667 (0.014) |
| GLMBoost | Glmnet | 0.649 (0.014) | 0.661 (0.018) | 0.641 (0.012) | 0.617 (0.010) | 0.679 (0.022) | 0.648 (0.013) | 0.647 (0.012) |
| GLMBoost | HDDA | 0.583 (0.038) | 0.580 (0.038) | 0.594 (0.036) | 0.575 (0.044) | 0.592 (0.040) | 0.571 (0.045) | 0.619 (0.037) |
| GLMBoost | LDA | 0.656 (0.019) | 0.648 (0.018) | 0.665 (0.020) | 0.684 (0.021) | 0.627 (0.017) | 0.655 (0.018) | 0.706 (0.016) |
| GLMBoost | LightGBM | 0.588 (0.013) | 0.594 (0.015) | 0.584 (0.012) | 0.557 (0.013) | 0.618 (0.018) | 0.587 (0.013) | 0.595 (0.021) |
| GLMBoost | Naïve Bayes | 0.661 (0.023) | 0.652 (0.026) | 0.673 (0.020) | 0.696 (0.013) | 0.626 (0.036) | 0.660 (0.023) | 0.693 (0.018) |
| GLMBoost | NNet | 0.505 (0.014) | 0.501 (0.016) | 0.507 (0.013) | 0.466 (0.019) | 0.539 (0.014) | 0.499 (0.016) | 0.546 (0.015) |
| GLMBoost | RF | 0.632 (0.016) | 0.636 (0.019) | 0.629 (0.016) | 0.623 (0.020) | 0.637 (0.028) | 0.630 (0.016) | 0.648 (0.015) |
| GLMBoost | SVM | 0.648 (0.021) | 0.646 (0.024) | 0.651 (0.020) | 0.656 (0.017) | 0.639 (0.031) | 0.647 (0.022) | 0.689 (0.019) |
| GLMBoost | XGB | 0.568 (0.015) | 0.568 (0.014) | 0.569 (0.017) | 0.570 (0.021) | 0.566 (0.016) | 0.567 (0.015) | 0.607 (0.020) |
| RF | BayesGLM | 0.599 (0.010) | 0.597 (0.011) | 0.603 (0.010) | 0.611 (0.022) | 0.588 (0.029) | 0.599 (0.010) | 0.618 (0.013) |
| RF | GLMBoost | 0.637 (0.015) | 0.645 (0.020) | 0.633 (0.013) | 0.622 (0.022) | 0.653 (0.030) | 0.636 (0.015) | 0.651 (0.006) |
| RF | Glmnet | 0.669 (0.017) | 0.698 (0.017) | 0.650 (0.015) | 0.600 (0.023) | 0.727 (0.022) | **0.664 (0.021)** | 0.655 (0.012) |
| RF | HDDA | 0.620 (0.006) | 0.608 (0.004) | 0.642 (0.009) | 0.690 (0.016) | 0.549 (0.015) | 0.615 (0.006) | 0.626 (0.008) |
| RF | LDA | 0.603 (0.011) | 0.599 (0.015) | 0.608 (0.005) | 0.625 (0.024) | 0.581 (0.044) | 0.602 (0.012) | 0.618 (0.009) |
| RF | LightGBM | 0.607 (0.011) | 0.615 (0.017) | 0.601 (0.008) | 0.576 (0.014) | 0.637 (0.028) | 0.606 (0.011) | 0.644 (0.014) |
| RF | Naïve Bayes | 0.643 (0.014) | 0.627 (0.012) | 0.666 (0.017) | 0.711 (0.022) | 0.575 (0.022) | 0.641 (0.014) | 0.662 (0.010) |
| RF | NNet | 0.553 (0.013) | 0.554 (0.011) | 0.553 (0.016) | 0.545 (0.027) | 0.560 (0.008) | 0.550 (0.014) | 0.576 (0.009) |
| RF | RF | 0.616 (0.006) | 0.619 (0.009) | 0.614 (0.005) | 0.604 (0.016) | 0.626 (0.020) | 0.615 (0.006) | 0.680 (0.010) |
| RF | SVM | 0.562 (0.024) | 0.564 (0.026) | 0.562 (0.023) | 0.559 (0.036) | 0.563 (0.047) | 0.559 (0.025) | 0.583 (0.014) |
| RF | XGB | 0.607 (0.016) | 0.612 (0.017) | 0.603 (0.016) | 0.591 (0.028) | 0.623 (0.025) | 0.606 (0.016) | 0.650 (0.030) |

**Supplementary Table S17. A comparison of prediction performance among different feature combinations in predicting rOA initial transition within 48 months.** Only the one among the 22 predictive models with the highest F1 score was shown for each feature combination. Models were evaluated through the accuracy (ACC), F1 score, AUC, recall, precision and their standard deviations. "RF" indicates performing feature selection based on their random forest permutation importance; "GLMBoost" means performing feature selection based on GLMBoost importance. The optimal F1 and AUC were highlighted in bold texts.

| **Test dataset** | | | | | | | | | |
|---|---|---|---|---|---|---|---|---|---|
| **Feature Combination** | **Feature Selection** | **Baseline Model** | **ACC** | **Precision (KLG = 2)** | **Precision (KLG < 2)** | **Recall (KLG = 2)** | **Recall (KLG < 2)** | **F1** | **AUC** |
| Demographic Raw Centile | RF | Glmnet | 0.641 (0.032) | 0.664 (0.030) | 0.625 (0.033) | 0.570 (0.056) | 0.674 (0.046) | 0.630 (0.040) | 0.628 (0.026) |
| Demographic Raw | GLMBoost | LightGBM | 0.596 (0.033) | 0.593 (0.035) | 0.599 (0.030) | 0.617 (0.019) | 0.574 (0.059) | 0.595 (0.034) | 0.627 (0.043) |
| Raw | GLMBoost | Naïve Bayes | 0.587 (0.017) | 0.565 (0.013) | 0.630 (0.027) | 0.752 (0.025) | 0.422 (0.019) | 0.575 (0.017) | 0.593 (0.041) |
| Raw Centile | RF | Glmnet | 0.659 (0.027) | 0.679 (0.028) | 0.643 (0.028) | 0.600 (0.048) | 0.700 (0.047) | **0.653 (0.036)** | **0.639 (0.021)** |
| **Validation dataset** | | | | | | | | | |
| **Feature Combination** | **Feature Selection** | **Baseline Model** | **ACC** | **Precision (KLG = 2)** | **Precision (KLG < 2)** | **Recall (KLG = 2)** | **Recall (KLG < 2)** | **F1** | **AUC** |
| Demographic Raw Centile | GLMBoost | Naïve Bayes | 0.667 (0.017) | 0.658 (0.018) | 0.680 (0.016) | 0.702 (0.013) | 0.633 (0.023) | **0.666 (0.017)** | **0.696 (0.012)** |
| Demographic Raw | RF | Naïve Bayes | 0.626 (0.008) | 0.603 (0.007) | 0.665 (0.015) | 0.740 (0.023) | 0.512 (0.018) | 0.621 (0.007) | 0.678 (0.013) |
| Raw | GLMBoost | LDA | 0.629 (0.012) | 0.620 (0.009) | 0.639 (0.015) | 0.664 (0.021) | 0.592 (0.008) | 0.628 (0.012) | 0.668 (0.014) |
| Raw Centile | RF | Glmnet | 0.669 (0.017) | 0.698 (0.017) | 0.650 (0.015) | 0.600 (0.023) | 0.727 (0.022) | 0.664 (0.021) | 0.655 (0.012) |

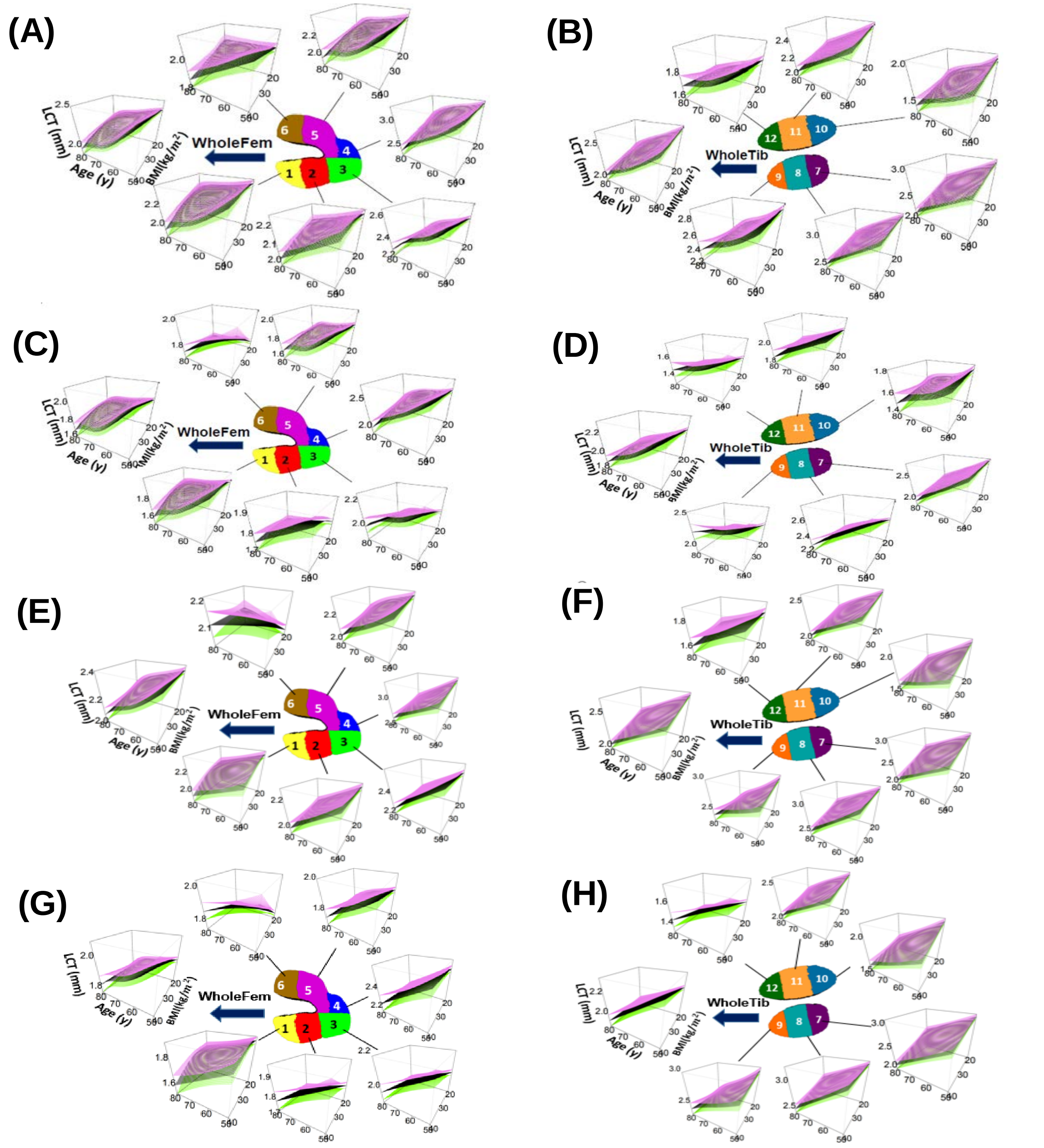


**Supplementary Figure S1. Three-dimensional perspective plots of the constructed ROI-level FKTR reference charts for (BMI, age) trajectories.** The fixed effect of 2-dimensional nonlinear BMI-age interactions within 14 knee cartilage regions were shown for the right (panels A, B, C and D) and left knee (panels E, F, G and H), respectively, while sex, race, height, and age-sex and BMI-sex interactions were controlled. Panels A, B, E, and F were for males while C, D, G and H were for females; A, C, E, and G were femoral regions while B, D, F and H were for tibial regions.

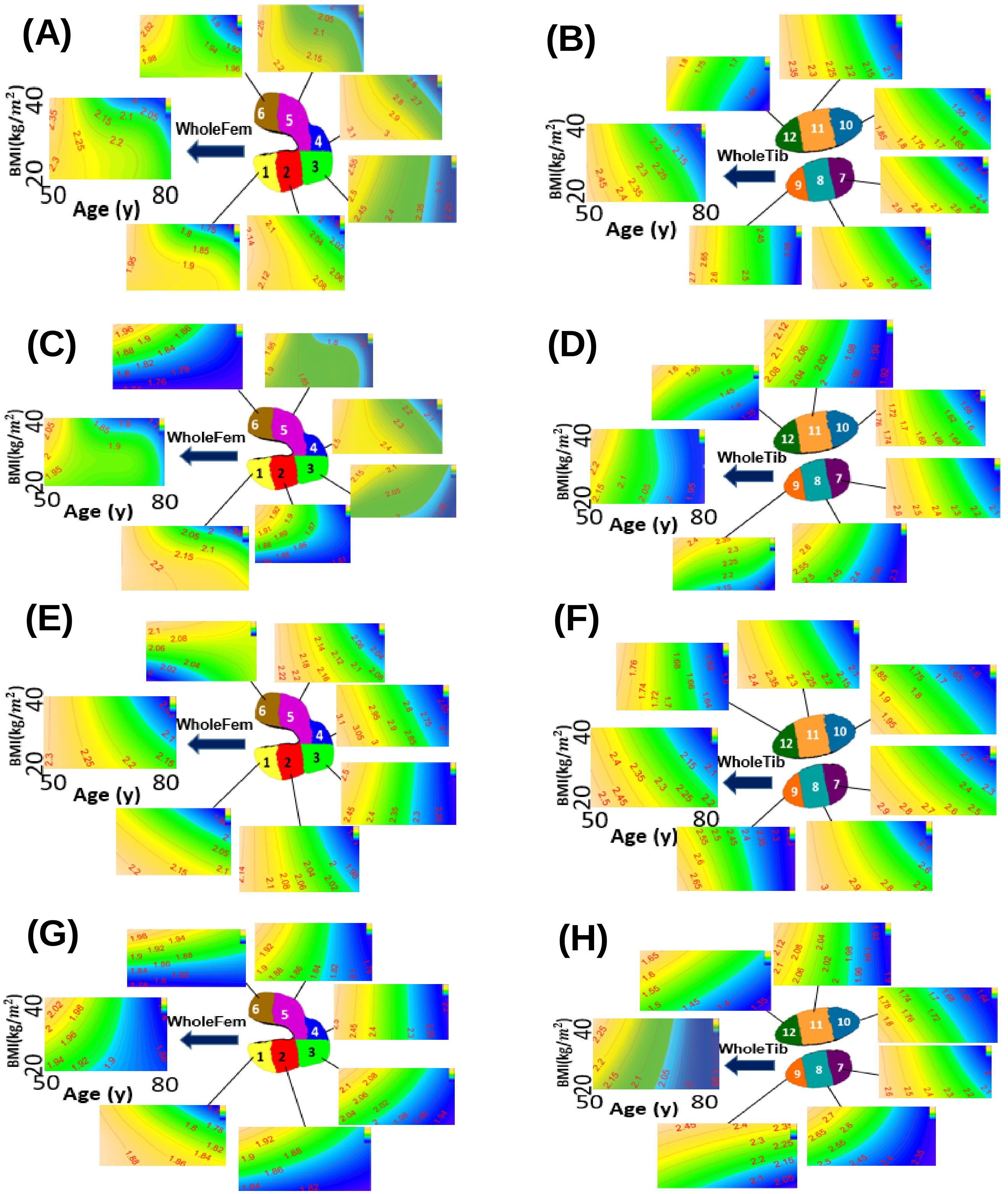


**Supplementary Figure S2. Two-dimensional contour plots of the constructed ROI-level FKTR reference charts for (BMI, age) trajectories.** Fixed effect of 2-dimensional nonlinear BMI-age interactions within 12 knee cartilage regions were shown for the right (panels A, B, C and D) and left knee (panels E, F, G and H), respectively, while sex, race, height, and age-sex and BMI-sex interactions were controlled as confounding factors. Panels A, B, E, and F were for males while C, D, G and H were for females; A, C, E, and G were femoral regions while B, D, F and H were for tibial regions.

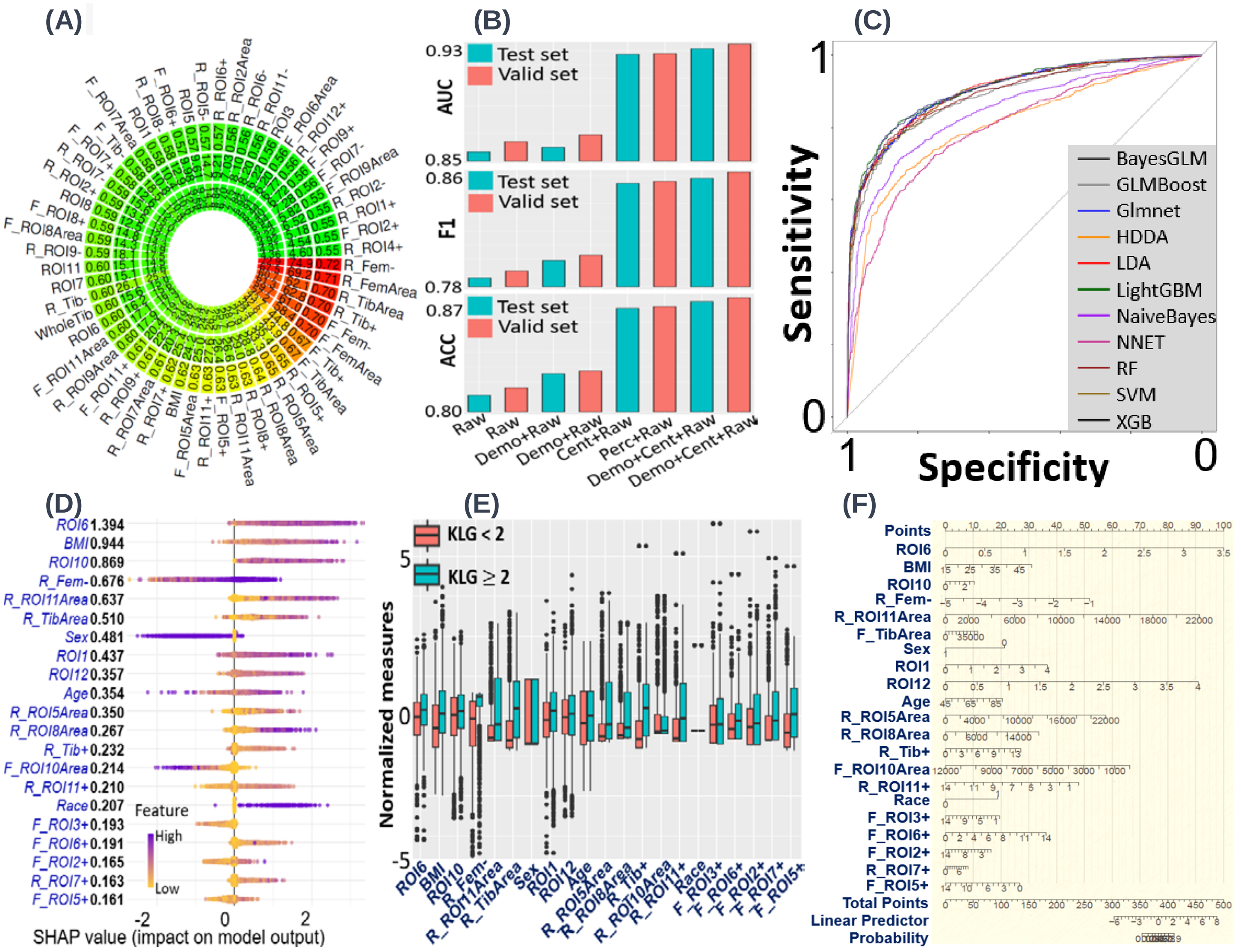


**Supplementary Figure S3. Results of rOA classification between KLG < 2 and KLG ≥ 2. (A)** Circular plot showing associations between radiographic OA (rOA) status KLG ≥ 2 and top predictors. Outer, middle, and inner circles signify Area Under the Receiver Operating Characteristic Curve (AUC), -Log10*p*, and -Log10(Bonferroni-adjusted *p*), respectively. Only results with AUC > *0.55* are shown. **(B)** Barplots illustrating the median performance metrics (AUC, F1 score, Accuracy) across *22* machine learning methods using different predictor combinations ("Demo" for demographic factors, "Raw" for raw regional cartilage thicknesses and "Cent" for centile features). Performance for test (blue) and validation (red) datasets is indicated. **(C)** Receiver Operating Characteristic (ROC) curves of the *11* models (each using Glmboost for variable selection) on the test set. The LightGBM model achieves the highest AUC and F1. **(D)** SHapley Additive exPlanations (SHAP) plot revealing impacts of top 21 predictors (> 80% contribution) on the test set based on the LightGBM model. Predictors are sorted by their importance; for each predictor, each dot represents a patient's SHAP value plotted horizontally, colored by the value of the feature from low (yellow) to high (purple). If purple and yellow dots are plotted at the lower and higher side, respectively, the KLG ≥ 2 risk becomes higher as the feature increases. **(E)** Boxplots of the top predictors (normalized to mean zero and unit variance) for KLG < 2 (red) and KLG ≥ 2 (blue). **(F)** Nomogram depicting coefficients of top predictors in a joint logistic model. The x-axis ("points") refers to the scores assigned to predictors based on their influence or weight in the predictive model.

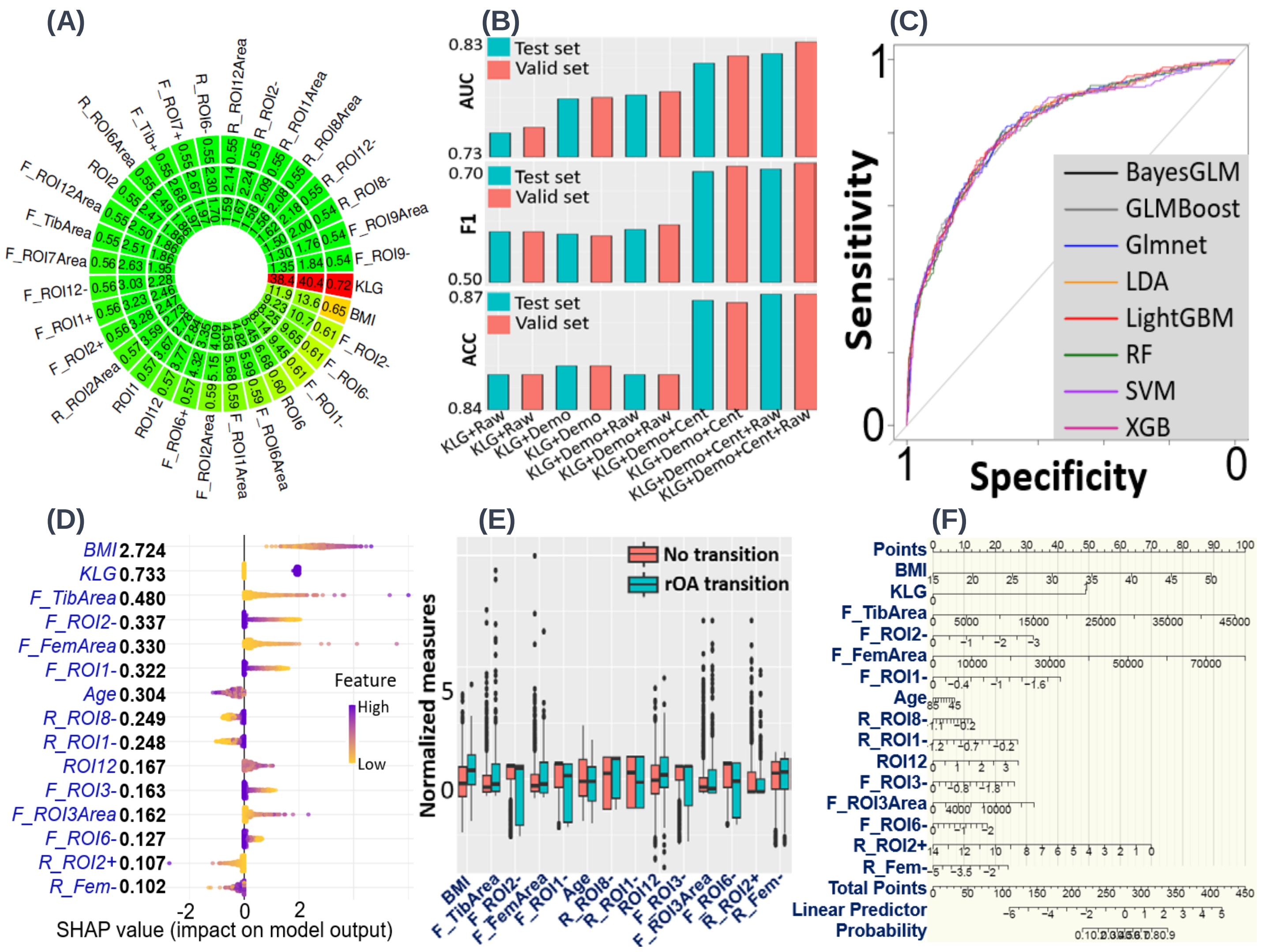

**Supplementary Figure S4. Results of incidence rOA prediction. (A)** Circular plot showing associations between future rOA incidence and predictors including raw cartilage thickness and centile features. Outer, middle, and inner circles signify Area Under the Receiver Operating Characteristic Curve (AUC), -Log10*p*, and -Log10(False Discovery Rate-adjusted *p*), respectively. Only associations with adjusted $p < 0.05$ are shown. **(B)** Barplots illustrating the median performance metrics (AUC, F1 score, Accuracy) across *22* machine learning methods using different predictor combinations (current Kellgren-Lawrence grading (KLG) score, demographic factors, raw regional cartilage thicknesses and centile features). Performance for test (blue) and validation (red) datasets is indicated. **(C)** Receiver Operating Characteristic (ROC) curves of *8* top models (each using GLMBoost for variable selection) on the test set. **(D)** SHapley Additive exPlanations (SHAP) plot revealing impacts of top predictors on the test set based on the LDA model. Predictors are sorted by their importance; for each predictor, each dot represents a patient's SHAP value plotted horizontally, colored by the value of the feature from low (yellow) to high (purple). If purple and yellow dots are plotted at the lower and higher side, respectively, the rOA incidence risk becomes higher as the feature increases. **(E)** Boxplots of the top predictors (normalized to mean zero and unit variance) for rOA transition (blue) and no transition (red). **(F)** Nomogram depicting coefficients of top predictors based on logistic model.

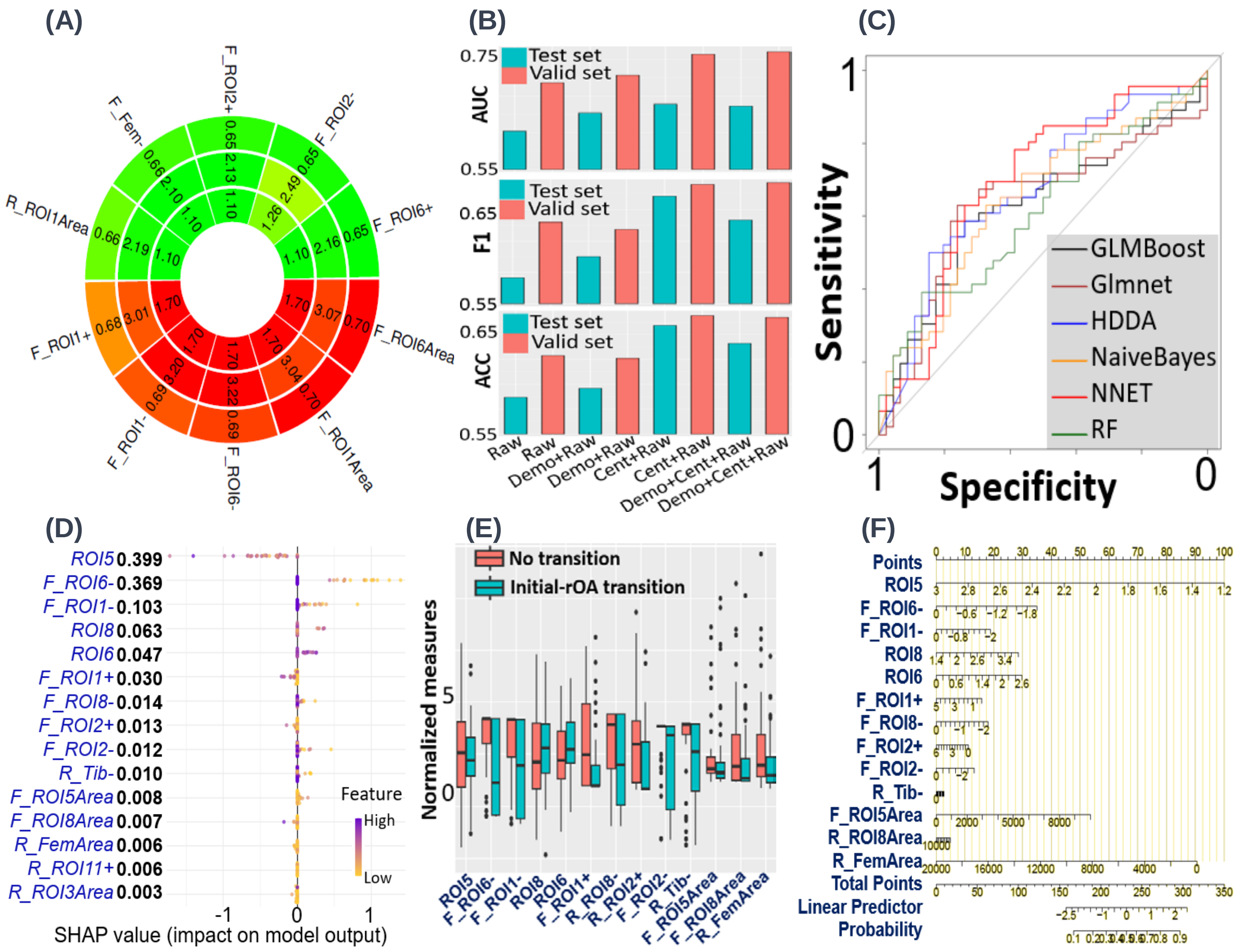

**Supplementary Figure S5. Results of initial-rOA transition prediction. (A)** Circular plot showing associations between initial-rOA transition and predictors including raw cartilage thickness and centile features. Outer, middle, and inner circles signify Area Under the Receiver Operating Characteristic Curve (AUC), -Log10*p*, and -Log10(Bonferroni-adjusted *p*), respectively. Only associations with adjusted $p < 0.05$ are shown. **(B)** Barplots illustrating the median performance metrics (AUC, F1 score, Accuracy) across *22* machine learning methods using different predictor combinations (demographic factors, raw regional cartilage thicknesses and centile features). Performance for test (blue) and validation (red) datasets is indicated. **(C)** Receiver Operating Characteristic (ROC) curves of *6* top models (each using random forest (RF) for variable selection) on the test set. **(D)** SHapley Additive exPlanations (SHAP) plot revealing impacts of top predictors on the test set based on the optimal model. Predictors are sorted by their importance; for each predictor, each dot represents a patient's SHAP value plotted horizontally, colored by the value of the feature from low (yellow) to high (purple). If purple and yellow dots are plotted at the lower and higher side, respectively, the initial-rOA transition risk becomes higher as the feature increases. **(E)** Boxplots of the top predictors (normalized to mean zero and unit variance) for with (blue) and without (red) rOA initial transition. **(F)** Nomogram depicting coefficients of top predictors based on logistic model with the elastic net penalty.
